# Towards a differential diagnosis of cochlear synaptopathy and outer-hair-cell deficits in mixed sensorineural hearing loss pathologies

**DOI:** 10.1101/19008680

**Authors:** Viacheslav Vasilkov, Sarah Verhulst

## Abstract

Damage to the auditory periphery is more widespread than predicted by the gold-standard clinical audiogram. Noise exposure, ototoxicity and aging can destroy cochlear inner-hair-cell afferent synapses and result in a degraded subcortical representation of sound while leaving hearing thresholds unaffected. Damaged afferent synapses, i.e. cochlear synaptopathy, can be quantified using histology, but a differential diagnosis in living humans is difficult: histology cannot be applied and existing auditory evoked potential (AEP) metrics for synaptopathy become insensitive when other sensorineural hearing impairments co-exist (e.g., outer-hair-cell damage associated with elevated hearing thresholds). To develop a non-invasive diagnostic method which quantifies synaptopathy in humans and animals with normal *or* elevated hearing thresholds, we employ a computational model approach in combination with human AEP and psychoacoustics. We propose the use of a sensorineural hearing loss (SNHL) map which comprises two relative AEP-based metrics to quantify the respective degrees of synaptopathy and OHC damage and evaluate to which degree our predictions of AEP alterations can explain individual data-points in recorded SNHL maps from male and female listeners with normal or elevated audiometric thresholds. We conclude that SNHL maps can offer a more precise diagnostic tool than existing AEP methods for individual assessment of the synaptopathy *and* OHC-damage aspect of sensorineural hearing loss.

**Significance Statement:** Hearing loss ranks fourth in global causes for disability and risk factors include noise exposure, ototoxicity and aging. The most vulnerable parts of the cochlea are the inner-hair-cell afferent synapses and their damage (cochlear synaptopathy) results in a degraded subcortical representation of sound. While synaptopathy can be estimated reliably using histology, it cannot be quantified this way in living humans. Secondly, other co-existing sensorineural hearing deficits (e.g., outer-hair-cell damage) can complicate a differential diagnosis. To quantify synaptopathy in humans and animals with normal or elevated hearing thresholds, we adopt a theoretical and interdisciplinary approach. Sensitive diagnostic metrics for synaptopathy are crucial to assess its prevalence in humans, study its impact on sound perception and yield effective hearing restoration strategies.

## Introduction

Inner-hair-cell (IHC) afferent synapses are the most vulnerable structures in the cochlea as they can be permanently damaged after noise exposure, ageing and ototoxicity while leaving the hair-cell bodies and mechanoreceptors intact, resulting in cochlear synaptopathy (Kujawa and Liberman, 2009; Bourien et al., 2014; Parthasarathy and Kujawa, 2018). Since its discovery, cochlear synaptopathy has been confirmed in a range of species (mice, guinea pigs, gerbils, rats, rhesus monkeys; Kujawa and Liberman, 2009; Furman et al., 2013; Bourien et al., 2014; Möhrle et al., 2016; Suzuki et al., 2016; Valero et al., 2017) including humans (Makary et al., 2011; Viana et al., 2015; Wu et al., 2018). However, its prevalence in humans and role for sound perception remain unclear. Synaptopathy does not affect objective hearing-threshold measures derived from ear-canal-recorded distortion-product otoacoustic emissions (DPOAEs; Trautwein et al., 1996) or scalp-electrode-recorded auditory evoked potentials (AEPs; Kujawa and Liberman, 2009). By extension, the behavioral pure-tone audiogram is expected to be insensitive to synaptopathy as objective and perceptual hearing threshold metrics correlate well (Van der Drift et al., 1987; Boege and Janssen, 2002). The insensitivity of threshold measures to synaptopathy can explain why synaptopathy is left undiagnosed in clinical practise and why its role in degrading sound perception for people with normal audiograms (“Hidden Hearing Loss”; Schaette and McAlpine, 2011; Plack et al., 2014) remains unclear. Because the onset of age-induced synaptopathy occurs prior to outer-hair-cell (OHC) damage associated with elevated hearing thresholds (e.g., in mice or in human post-mortem material Sergeyenko et al., 2013; Parthasarathy and Kujawa, 2018; Makary et al., 2011; Viana et al., 2015), the prevalence of synaptopathy in humans is expected to be high, especially because abnormal audiograms are a normal consequence of the human aging process (ISO 7029).

The development of a non-invasive diagnostic test for synaptopathy in humans is both necessary and urgent, and several studies have proposed methods on the basis of auditory brainstem response (ABR) or envelope-following response (EFR) alterations observed in animals with synaptopathy. The ABR is a transient-evoked AEP of which the first five wave peaks reflect synchronous aggregate neural information from ascending peripheral auditory processing centers (Picton, 2010) and reduced ABR wave-I amplitudes reflect the degree of histological synaptopathy in animal models for stimulus levels above the hearing threshold (Kujawa and Liberman, 2009; Furman et al., 2013; Möhrle et al., 2016). The EFR is an AEP to sustained stimulation with amplitude-modulated sound and its magnitude reflects neuronal population coding to the stimulus envelope. For modulation frequencies above 80 Hz, the sources of the EFR have been assigned to subcortical neuronal populations in the auditory-nerve (AN), cochlear nucleus (CN) and inferior colliculus (IC) (Dolphin and Mountain, 1992; Kuwada et al., 2002; Purcell et al., 2004; Bidelman, 2015). Several studies have shown that the EFR is compromised by synaptopathy (for modulation frequencies between 700 and 1000 Hz; Shaheen et al., 2015; Parthasarathy and Kujawa, 2018), and that its magnitude correlates to performance in a behavioral temporal envelope detection task in listeners with normal audiograms (Bharadwaj et al., 2015). The EFR might thus be a good candidate for synaptopathy diagnosis in humans. However, a direct translation of animal-based AEP metrics to human synaptopathy diagnostics has proven difficult because a direct quantification of synaptopathy is only possible via histology and hence impossible in living humans. Secondly, inter-individual differences in neural background-noise level, sex and head size (Trune et al., 1988; Mitchell et al., 1989; Hickox et al., 2017) can confound the interpretation of AEP amplitudes in terms of hearing status and require differential AEP metrics (e.g., Bharadwaj et al., 2015; Verhulst et al., 2016; Guest et al., 2018; Bramhall et al., 2019). Lastly, AEP metrics are not only compromised by synaptopathy, OHC damage is also known to affect the AEP (e.g., Gorga et al., 1985; Herdman and Stapells, 2003; Chen et al., 2007; Garrett and Verhulst, 2019). This renders any interpretation of AEP amplitudes in terms of synaptopathy impossible without controlling for the degree of OHC damage as well.

As OHC damage is common in humans, and existing AEP-based metrics for synaptopathy assessment have so far mostly focused on animals and humans with normal hearing thresholds, it is crucial to understand to which degree co-existing OHC damage affects the AEP metrics and compromises their diagnostic sensitivity. To this end, this study adopts an interdisciplinary approach which combines computational modeling with human AEP recordings to study how synaptopathy and OHC damage affect AEP metrics commonly used in synaptopathy studies. On the basis of these results, we propose the use of a *sensorineural hearing loss (SNHL) map* which comprises two relative AEP metrics to maximally separate and quantify the synaptopathy aspect of hearing loss. Modeling can surpass experimental limitations in human studies and offers a means to transfer insight from animal histology to human applications with a number of advantages: (i) the respective impact of each impairment (OHC and different AN fiber types) on the AEP can be studied separately and in combination, which is difficult to establish experimentally as this would require different cochlear damage manipulations in a single animal. (ii) Models can connect changes in single-unit physiology to its alteration in population AEP responses directly, which is experimentally challenging. Lastly (iii), because human models of cochlear mechanics can be adopted to simulate SNHL and AEPs, the study results can directly be applied to humans and hence avoid experimental inter-species AEP differences (Hickox et al., 2017). We first adopt a modelling approach to study how different types of SNHL affect the AEP to basic stimuli applied in synaptopathy studies, after which we evaluate how AEPs can be used to diagnose synaptopathy in people with normal *or* abnormal audiograms. We focus this study on AEP metrics used in clinical practice (the ABR) and those adopted in several synaptopathy studies in subjects/animals with normal hearing sensitivity (the EFR; Bharadwaj et al., 2015; Shaheen et al., 2015; Guest et al., 2018; Parthasarathy and Kujawa, 2018). This approach maximizes the clinical uptake of our methods, while allowing for a comparison of our model predictions with existing animal synaptopathy data and human reference recordings.

## Materials and Methods

### Computational model of the human auditory periphery

The auditory periphery model (Fig.1) we adopted for this study accurately captures human aspects of cochlear filter tuning, the level-dependence of ABR and EFR responses, and can simulate hearing impairments related to OHC damage and synaptopathy (Verhulst et al., 2018, model implementation v1.2). The model simulates how the ascending auditory pathway processes acoustic input, and includes middle-ear filtering, a nonlinear transmission-line representation of human cochlear mechanics (Verhulst et al., 2012; Altoè et al., 2014), a biophysical model of the IHC-AN complex (Altoè et al., 2018), as well as a phenomenological description of ventral cochlear nucleus (CN) and inferior colliculus (IC) neurons (Nelson and Carney, 2004). The model is suited for the purpose of this study as it reasonably captures how OHC damage and synaptopathy affect human ABRs and EFRs for different sound intensities (Verhulst et al., 2016, 2018). Additionally, because the model includes a transmission-line description of the cochlear mechanics (Zweig, 1991; Altoè et al., 2014), it simulates human otoacoustic emissions (OAEs), which were used to calibrate the tuning characteristics of the simulated cochlear filters across the cochlear partition (Shera et al., 2010; Verhulst et al., 2012).

**Figure 1.**
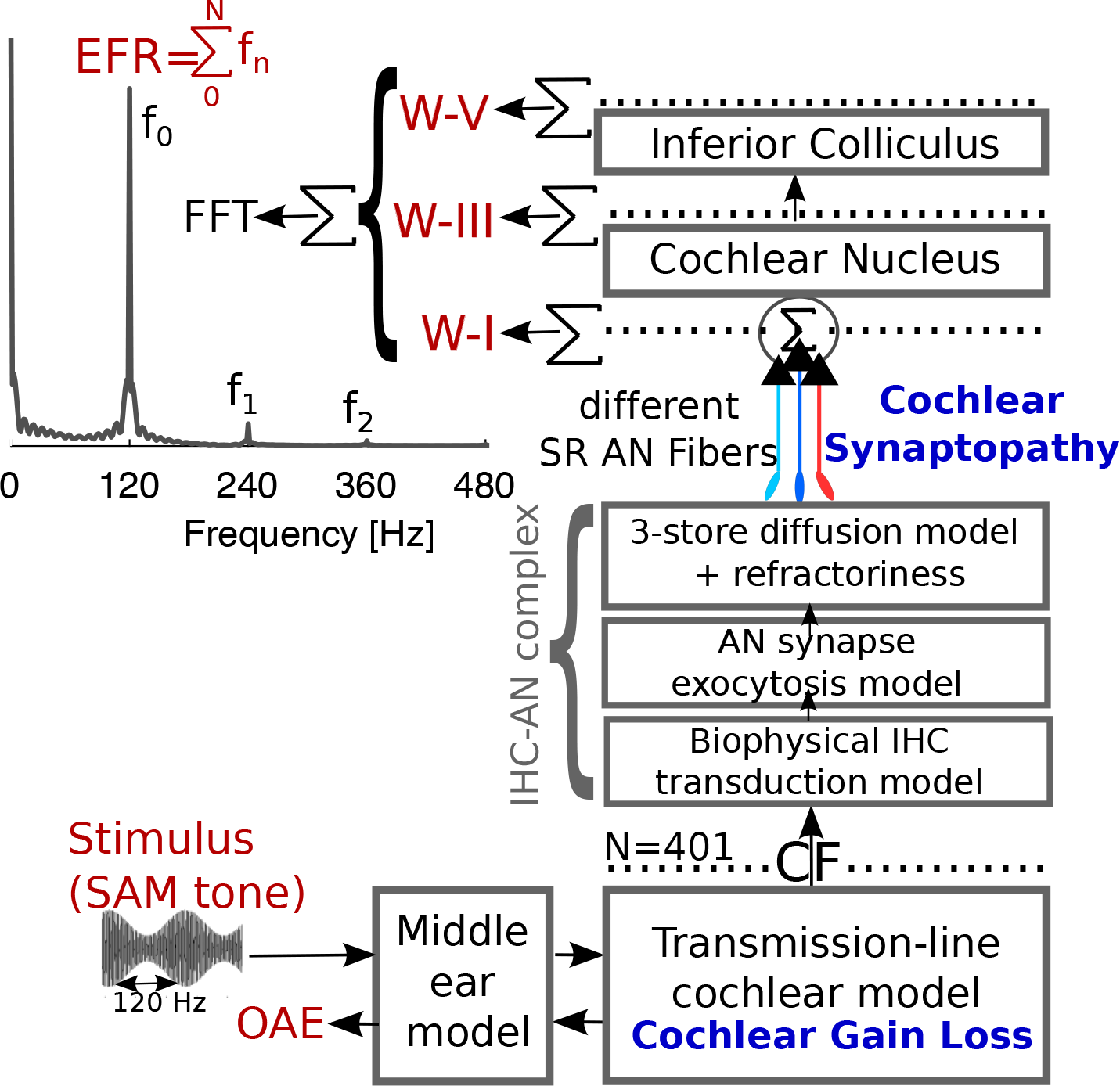
An overview of the computational model of the human auditory periphery (Verhulst et al., 2018) which was used to simulate ABR and EFRs. Acoustic stimuli pass through a middle-ear and transmission-line cochlear model which computes the BM velocity over N=401 CFs spaced between 112 Hz and 12 kHz along the cochlear partition. At each CF, BM velocity passes through a biophysical model of the IHC-AN complex (Altoè et al., 2018) which simulates responses for 3 LSR, 3 MSR and 13 HSR fibers, whose responses are summed and passed to a functional model of the CN and IC (Nelson and Carney, 2004). ABR W-I and W-V were simulated by adding up responses across the simulated CFs at the AN and IC layers, respectively. The EFR was obtained by summing up activity across the same CFs and three layers of processing (i.e., at the output of AN, CN and IC) after which an FFT magnitude spectrum was computed to derive the EFR metrics.

#### Simulating outer-hair-cell damage

The adopted cochlear mechanics model allows for a straightforward implementation of frequency-specific OHC damage (Verhulst et al., 2016, 2018) caused by damaged mechano-receptors or presbycusis associated with wider cochlear filters and reduced hearing sensitivity. OHC damage was simulated by reducing cochlear filter gain relative to the models’ normal-hearing filter gain (in dB HL) at low stimulus levels and at characteristic frequencies (CFs) corresponding to the audiometric testing frequencies. The relationship between cochlear filter gain and the value of the double-pole of the cochlear admittance of the model is described in Verhulst et al. (2016) and Fig. 2a (left) shows examples of the different sloping high-frequency cochlear gain loss profiles considered here. In analogy to the behavioral audiogram, the models’ detection sensitivity is reduced by an amount corresponding to the applied dB HL filter gain reduction. We limited our simulations to a range of common sloping high-frequency gain loss profiles characterized by the dB HL loss difference between 1 and 4-kHz and included one sloping profile with a normal gain up to 4-kHz, but a 35 dB HL loss at 8-kHz to simulate extended high-frequency (EHF) hearing loss.

**Figure 2.**
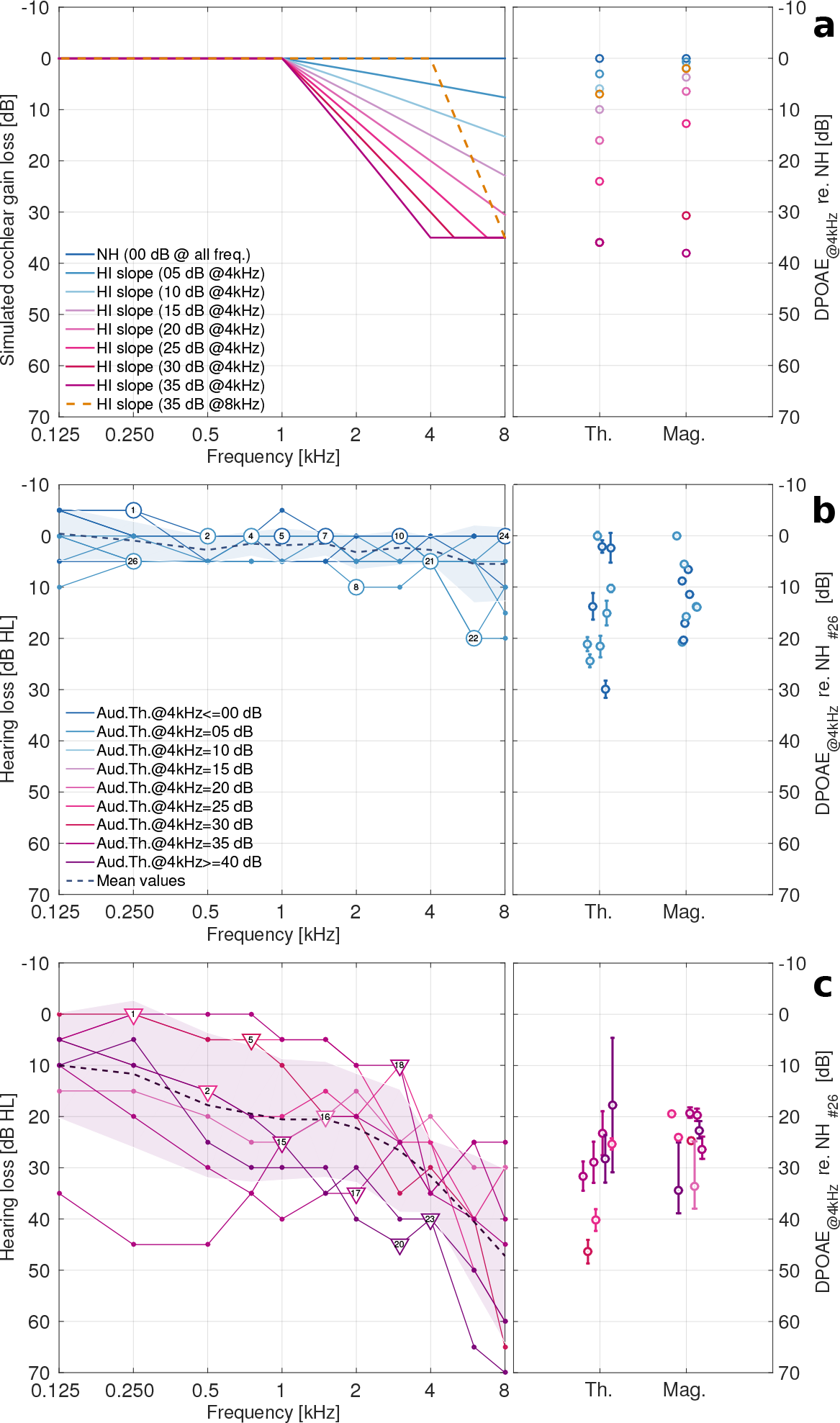
**Left: a** Cochlear gain loss profiles, which were used to simulate OHC damage. The degree of cochlear filter gain reduction is indicated using a dark blue to dark red color gradient and is labeled according to the dB HL values at 4-kHz. One sloping hearing loss profile with a 35 dB HL at 8-kHz was also considered (orange). **b** Measured pure-tone audiograms for 11 normal-hearing listeners (blue) and **c** 9 listeners with elevated high-frequency audiometric thresholds (red) who participated in both the SAM-tone AEP and behavioral AM DT experiments. Individual listeners are labeled can be followed across experiments. **Right: a** Simulated and **b,c** reference DPOAE threshold (**Th**.) and supra-threshold DPOAE magnitude **(Mag**.) shifts relative to the model NH profile or the best measured DPOAE (NH subject #26), respectively.

The cochlear gain loss profiles depicted in Fig. 2a only indirectly correspond to human audiograms as changes in behavioral detection thresholds were not simulated. However, a direct comparison between the OHC integrity of the model and that of the study participants is possible through DPOAE thresholds and their alterations as a function of the cochlear gain loss profile (models) or audiogram (humans). Reference DPOAEs were recorded from study participants with audiograms shown in Fig. 2b,c (left) who were separated into two groups (normal-hearing: NH and hearing-impaired: HI) on the basis of a better (Fig. 2b) or worse than 20 dB HL (Fig. 2c) 4-kHz audiometric threshold. DPOAE thresholds were derived from recorded DPOAE level series for an f_2_ of 4 kHz and a f_2_/f_1_ ratio of 1.2. L_2_ levels ranged in 6-dB steps between 24-66 dB for NH and 30-72 dB for HI participants and L_1_ levels were set according to the L_1_=44 + 0.45 L_2_ level paradigm (Neely et al., 2005). DPOAE thresholds (Fig. 2b,c; right) were determined as the L_2_ level at which a cubic fitting function through the data-points reached a level of −25dB SPL (human; Boege and Janssen, 2002) or 0 dB SPL (model). Additional details on the human experimental procedures and fitting function are given in Verhulst et al. (2016). The right panel of Fig. 2a depicts simulated DPOAE threshold shifts along with supra-threshold DPOAE level reductions (L_2_ of 70 dB) relative to the NH model, and shows that both simulated DPOAE thresholds and levels decrease by up to 40 dB as the cochlear gain loss amount increases. Human DPOAE thresholds and level differences were computed relatively to the strongest measured DPOAE, and fell within the range of simulated DPOAE shifts. We hence conclude that the simulated cochlear gain loss profiles capture a variety of OHC deficits and associated DPOAE reductions which corroborates DPOAE observations in human subjects with audiograms in Figs. 2b&c.

#### Simulating cochlear synaptopathy

Cochlear synaptopathy was modeled by selectively reducing the number of AN fibers of different types that synapse onto single IHC at each simulated tonotopic location. The NH model had 19 fibers with different spontaneous rates synapsing onto each IHC: 3 low (LSR), 3 medium (MSR) and 13 high (HSR) fibers, following the empirical ratio observed in cats (Liberman, 1978). Four synaptopathy profiles were implemented by removing the following fiber types across the tonotopic axis: (i) all LSR fibers (HI synapt_0L,3M,13H_), (ii) all LSR and MSR fibers (HI synapt_0L,0M,13H_), (iii) all LSR, MSR and 50 % of the HSR fibers (HI synapt_0L,0M,07H_), and (iv) all LSR, MSR and 80 % of the HSR fibers (HI synapt_0L,0M,03H_). We limited our simulations to uniform CF-independent synaptopathy profiles akin to the assumption of using a broadband stimulus to evoke cochlear synaptopathy. Aside from synaptopathy, no other IHC-specific dysfunctions were simulated as synaptopathy was shown to occur without destroying the sensory cells in several studies (Kujawa and Liberman, 2009; Lin et al., 2011; Furman et al., 2013; Shaheen et al., 2015).

#### Simulating evoked potentials

Capturing a variety of AN fiber types and their respective adaptation properties is essential for the simulation of EFRs to sustained amplitude-modulated stimuli (Verhulst et al., 2018). To this end, we adopted an IHC-AN synapse model which realistically captures IHC saturation and AN firing rate properties in response to pure tones and sinusoidally amplitude-modulated (SAM) tones of increasing stimulus intensity (Altoè et al., 2018). At each tonotopic location, instantaneous firing rates from 19 AN fibers of three SR types which synapse onto a single IHC were summed and projected to a single CN unit of the same CF. The postsynaptic activity of a single CN unit served as an input to a single IC unit. A same-frequency inhibition and excitation model for CN and IC units which captures the modulation filtering and onset enhancement characteristics of auditory brainstem and midbrain neurons was adopted (Nelson and Carney, 2004). Population responses were simulated by adding up activity across a tonotopic array of 401 IHC-AN/CN/IC units with CFs between 112 Hz and 12 kHz (distributed according to the frequency-position map Greenwood, 1990) at three processing stages: the AN (after summing up 19 AN fibers across each IHC with different CFs) which yields the W-I response in Fig. 1, and the CN and IC model stages yielding the W-III and W-V response respectively. Each simulated ABR wave peak (i.e., wave-I, AN activity; wave-III, CN activity; wave-V, IC activity) was scaled in amplitude according to NH human reference ABR data (Table 8-1 in Picton, 2010; Verhulst et al., 2018) and ABR waves were treated separately as their peaks did not overlap in time. For EFR simulations, the scaled population responses from the AN, CN and IC stage were added up to capture the different subcortical sources that contribute to the EFR (Dolphin and Mountain, 1992; Kuwada et al., 2002).

### Evoked potential recordings and data analysis

AEPs were recorded from a total of 46 participants with audiometrically normal-hearing (NH; n=25, age: 25.9 *±* 4.3, 16 females) and hearing-impaired (HI; n=21, age: 66.2 *±* 7.6, 9 females) thresholds. HI listeners were recruited on the basis of a sloping high-frequency audiogram that exceeded 20 dB HL at 4-kHz. For visual clarity, Figure 2b,c shows individual audiograms of a subset of participants for whom ABRs, EFRs, DPOAEs and psychoacoustic thresholds were collected within one of two experimental sessions. The audiograms were devised in NH (Fig. 2b, n=11, age: 26.7 *±* 3.9, 7 females) and HI (Fig. 2c, losses *>* 20 dB HL, n=9, age: 70.9 *±* 5.5, 3 females) groups. The audiometric thresholds, gender and ages of all listeners who participated in the ABR and EFR experiments within both PT and BB sessions (i.e., with EFRs evoked by SAM pure tone and broadband noise, respectively), but not necessarily in the psychoacoustic experiment, are listed in Table I. Participants received written and oral information about the experiments, and protocols were approved by the ethics commission of Oldenburg University. Subjects volunteered for the study, gave a written informed consent, and were paid for their participation.

**Table I.** Participant profiles. Gender (G), age (A, years), and pure-tone audiometric thresholds (dB HL) for the tested ear and audiometric testing frequencies as well as the pure tone average (PTA, dB HL) across presented frequencies (kHz).

| Subj# | G | A | 0.125 | 0.25 | 0.5 | 0.75 | 1 | 1.5 | 2 | 3 | 4 | 6 | 8 | PTA |
| --- | --- | --- | --- | --- | --- | --- | --- | --- | --- | --- | --- | --- | --- | --- |
| Audiometrically normal hearing group |  |  |  |  |  |  |  |  |  |  |  |  |  |  |
| 1 | m | 26.7 | -5 | -5 | 5 | 0 | 0 | 0 | 5 | 5 | 0 | 5 | 10 | 1.82 |
| 2 | f | 27.9 | -5 | 0 | 0 | 0 | 0 | 0 | 0 | 0 | 5 | 5 | 0 | 0.45 |
| 3 | f | 25.5 | 0 | 0 | 0 | 0 | -5 | 0 | 5 | 0 | 5 | 5 | 5 | 1.36 |
| 4 | f | 20.2 | 5 | 0 | 0 | 0 | 0 | 0 | 0 | 0 | 5 | 0 | 0 | 0.91 |
| 5 | m | 25.2 | -5 | -5 | 0 | 0 | 0 | 0 | 5 | 0 | 0 | 0 | 0 | -0.45 |
| 6 | f | 25.1 | 5 | 5 | 5 | 5 | 5 | 0 | 0 | 5 | 5 | 0 | 0 | 3.18 |
| 7 | f | 24 | -5 | 0 | 0 | 0 | -5 | 0 | 0 | 0 | 0 | 0 | 0 | -0.91 |
| 8 | f | 32.4 | 0 | 5 | 5 | 5 | 5 | 5 | 10 | 10 | 5 | 5 | 5 | 5.45 |
| 9 | m | 28.6 | -5 | 0 | 0 | 5 | 0 | 0 | 0 | 0 | 0 | 0 | 5 | 0.45 |
| 10 | f | 21.2 | -5 | 0 | 0 | 0 | 5 | 5 | 5 | 0 | 0 | 0 | 0 | 0.91 |
| 11 | f | 26.3 | -5 | -5 | -5 | 0 | 0 | 0 | 0 | 5 | 5 | 5 | 5 | 0.45 |
| 12 | f | 29.1 | 0 | 0 | 0 | -5 | 0 | 0 | 5 | 0 | 0 | 5 | 5 | 0.91 |
| 13 | f | 21.5 | -5 | 0 | 0 | 0 | 0 | 0 | 0 | 0 | 0 | 0 | 0 | -0.45 |
| 14 | m | 24.7 | 0 | -5 | 0 | 0 | 0 | -5 | 0 | 0 | 0 | 0 | 5 | -0.45 |
| 15 | f | 28.9 | 0 | 0 | -5 | 0 | 0 | 0 | 0 | 0 | 0 | 5 | 0 | 0.00 |
| 16 | f | 21.4 | 0 | 5 | 5 | 10 | 5 | 5 | 5 | 5 | 0 | 10 | 5 | 5.00 |
| 17 | f | 23.7 | -5 | 0 | 0 | 0 | 0 | 0 | 0 | 5 | 0 | 0 | 0 | 0.00 |
| 18 | f | 14.3 | 0 | 5 | 5 | 5 | 0 | 0 | 0 | 0 | 5 | 0 | 0 | 1.82 |
| 19 | m | 24.6 | -5 | -5 | 0 | 5 | 0 | 0 | 0 | 0 | 0 | 20 | 5 | 1.82 |
| 20 | m | 35.1 | 10 | 10 | 15 | - | 15 | - | 5 | - | 10 | 5 | 20 | 11.25 |
| 21 | f | 31.4 | 0 | 0 | 5 | - | 5 | - | 5 | 5 | 5 | 20 | 10 | 6.11 |
| 22 | m | 27.3 | 0 | 5 | 5 | 0 | 0 | 0 | 0 | 5 | 5 | 20 | 20 | 5.45 |
| 23 | m | 25.6 | 10 | 10 | 15 | 15 | 20 | 10 | 15 | 15 | 20 | 25 | 10 | 15.00 |
| 24 | f | 30.4 | 5 | 5 | 5 | 5 | 5 | 5 | 0 | 0 | 0 | 0 | 0 | 2.73 |
| 26 | m | 26.8 | 10 | 5 | 5 | 5 | 5 | 0 | 5 | 0 | 5 | 5 | 15 | 5.45 |
| Participants with impaired thresholds |  |  |  |  |  |  |  |  |  |  |  |  |  |  |
| 1 | m | 65 | 5 | 0 | 5 | 5 | 5 | 5 | 10 | 25 | 25 | 50 | 60 | 17.73 |
| 2 | f | 62.9 | 5 | 10 | 15 | 20 | 20 | 15 | 20 | 10 | 25 | 40 | 30 | 19.09 |
| 3 | m | 55.3 | 0 | 0 | 0 | - | 10 | - | 15 | 20 | 25 | 40 | 40 | 16.67 |
| 4 | m | 54.4 | 0 | 0 | 0 | - | 5 | - | 5 | 25 | 25 | 25 | 10 | 10.56 |
| 5 | f | 75.5 | 0 | 0 | 5 | 5 | 10 | 20 | 20 | 35 | 30 | 40 | 65 | 20.91 |
| 6 | f | 73.8 | 20 | 20 | 15 | 15 | 20 | 10 | 20 | 20 | 25 | 40 | 60 | 24.09 |
| 7 | f | 65 | 20 | 25 | 35 | 25 | 20 | 25 | 20 | 25 | 25 | 40 | 65 | 29.55 |
| 8 | f | 65.8 | 0 | 0 | 5 | 5 | 5 | 5 | 5 | 20 | 20 | 30 | 30 | 11.36 |
| 9 | f | 62.7 | -5 | 0 | 0 | 5 | 0 | 15 | 10 | 10 | 25 | 35 | 20 | 10.46 |
| 10 | f | 64.1 | 10 | 10 | 20 | 20 | 25 | 20 | 20 | 25 | 20 | 25 | 35 | 20.91 |
| 11 | f | 68.8 | 20 | 25 | 30 | 35 | 30 | 35 | 30 | 35 | 25 | 35 | 55 | 32.27 |
| 12 | m | 60.4 | 0 | 10 | 10 | 15 | 10 | 15 | 15 | 20 | 25 | 30 | 35 | 16.82 |
| 13 | m | 68.4 | 10 | 10 | 5 | 15 | 20 | 15 | 20 | 15 | 25 | 35 | 60 | 20.91 |
| 14 | m | 48.3 | 5 | 0 | 0 | 5 | 5 | 5 | 5 | 5 | 20 | 25 | 15 | 8.18 |
| 15 | m | 71.2 | 35 | 45 | 45 | 35 | 25 | 20 | 20 | 25 | 35 | 25 | 25 | 30.46 |
| 16 | m | 74.6 | 15 | 15 | 20 | 25 | 25 | 20 | 15 | 25 | 20 | 30 | 30 | 21.82 |
| 17 | m | 63.8 | 10 | 20 | 30 | 35 | 40 | 35 | 35 | 25 | 35 | 40 | 45 | 31.82 |
| 18 | m | 72.4 | 5 | 0 | 0 | 0 | 5 | 5 | 10 | 10 | 35 | 25 | 40 | 12.27 |
| 20 | f | 74.7 | 10 | 5 | 20 | 30 | 30 | 30 | 40 | 45 | 40 | 65 | 70 | 35.46 |
| 22 | m | 66.3 | 15 | 5 | 10 | 20 | 5 | 0 | 5 | 10 | 20 | 25 | 50 | 15.00 |
| 23 | m | 77.6 | 5 | 10 | 15 | 20 | 25 | 35 | 30 | 40 | 40 | 50 | 60 | 30.00 |

All stimuli were digitized with a sampling rate of 48 kHz for the AEP study and of 100 kHz for the model simulations. Both EFRs and ABRs were recorded in a single 1.5 hr recording session using a 32-channel EEG amplifier (BioSemi) with a sampling rate of 16384 Hz and a custom-built triggerbox. The electrodes were positioned equidistantly with center at Cz. Common-mode-sense and driven-right-leg (CMS/DRL) electrodes were placed on top of the head near Cz and two reference electrodes were placed on the earlobes. The Cz-channel potentials were re-referenced to the mean of the two earlobe electrode potentials to yield the AEP waveform. The Raw recordings are part of the data collection reported in Verhulst et al. (2016); Garrett and Verhulst (2019), which provide further detail on the recording configuration.

**ABR**s were recorded to 7000 repetitions of a 80-*µ*s condensation click presented monaurally with a rate of 33.3 Hz using a uniformly distributed 10% jitter on the recording window duration. Four stimulus levels were considered: 70, 80, 90, and 100 dB peSPL. ABR recordings were bandpass filtered between 200 Hz and 1.5 kHz, epoched within a −5 to 20 ms window relative to the stimulus onset, baseline corrected, epoch rejected (with 40 *µ*V threshold) and averaged using MATLAB. For each participant and stimulus condition, average ABR waveforms were estimated using a bootstrap procedure with 200 averages of 6000 randomly chosen epochs per estimate. ABR wave-V amplitudes [in *µ*V] were determined as the trough-to-peak difference between the global maximum (for 70 dB peSPL stimulus, within the [4-7.5] or [5-10] ms window for NH or HI listeners, respectively) and the waveform minimum which preceded the peak within 3 ms (Picton, 2010) or 4 ms window in NH and HI listeners, respectively). The longer window for HI listeners was applied to account for potentially less synchronized responses with increased peak latencies in this test group. This procedure was implemented as an automatic procedure and labeled peaks and troughs of the IV-V complex were visually inspected afterwards. No changes were made to automatically selected peaks and troughs after visual inspection (although HI subject #23 showed strong fluctuations before and after stimulus onset which could have affected the estimated ABR amplitudes). The standard deviation for ABR amplitudes was calculated using the propagation of error method based on the combination of deviations for the ABR wave-V maximum and minimum stemming from the 200 bootstrapped average waveforms.

**EFR**s were recorded in response to monaurally presented 600-ms long sinusoidally amplitude-modulated stimuli that were followed by a uniformly distributed random silence jitter (*>*90 and *<*110 ms; mean = 100 ms). The modulation depth was 0 dB re 100% modulation and the modulation-frequency was 120 Hz. Two carrier types were considered in the study: (i) a 4-kHz pure tone (PT condition; applied to 11 NH and 9 HI participants from the cohort; Fig. 2 b,c) and (ii) a white noise carrier with a 20-20000 Hz bandwidth (broadband (BB) condition, applied to 23 NH and 15 HI participants). Stimuli were ramped with a 5% tapered cosine (Tukey) window. Stimulus levels were 70 and 75 dB SPL for the PT and BB conditions, respectively. 800 stimulus repetitions (i.e., 400 of each polarity) were presented for the PT condition and 600 stimulus repetitions were presented for the BB condition. The earlobe-referenced EFR recordings were visually inspected to ensure that the Cz channel data was successfully recorded and eyeblink-artifacts were removed (further preprocessing details can be found in Garrett and Verhulst, 2019). The artifact-free data were 60-Hz high-pass and 650-Hz low-pass filtered using a 4^*th*^ order IIR Butterworth filter, epoched in 600-ms windows following stimulus onset, and baseline corrected. The EFR data were pre-processed using Python and MNE-Python scripts (Gramfort et al., 2013, 2014).

Average time-domain waveforms and EFR magnitudes as well as standard deviations and noise-floor estimates were obtained by implementing a bootstrapping method (Zhu et al., 2013) in MATLAB. Magnitude spectra (in *µ*V) were calculated from applying the fast (discrete) Fourier transform to the time-domain response of each bootstrapped average. The noise floor was calculated from 1000 averages of the total number of randomly sampled epochs (available after artifact rejection) in which half of the randomly drawn epochs were rotated in phase by 180^*°*^. Mean EFR magnitude and standard deviations were computed based on the magnitude spectra for 200 averages of randomly sampled epochs with replacement. Figure 3a shows an example of an averaged EFR magnitude spectrum and corresponding noise floor estimate for a NH listener in the cohort. For each average, spectral peaks (F_*n*_) at the stimulus modulation frequency (f_0_=120 Hz) and harmonics (f_(*k−*1)_=*k*\*f_0_, *k*=[1..5]) were identified, and corrected for by subtracting the respective estimated noise-floor value (NF_*n*_). The EFR magnitude was obtained after performing an inverse fast (discrete) Fourier transformation which included the noise-floor corrected peaks (F_*n*_ *−* NF_*n*_) and their corresponding phase angle values (*θ*_*n*_) to yield a time-domain signal which only contains energy at the modulation frequencies and harmonics. Energy at other frequencies were removed. The EFR magnitude was defined as half the peak-to-peak amplitude of the reconstructed time domain-signal waveform:

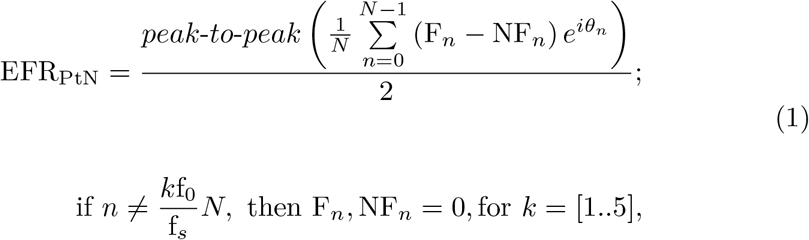

where *N* corresponds to the length of the magnitude spectrum, and f_*s*_ is the sampling rate.

**Figure 3.**
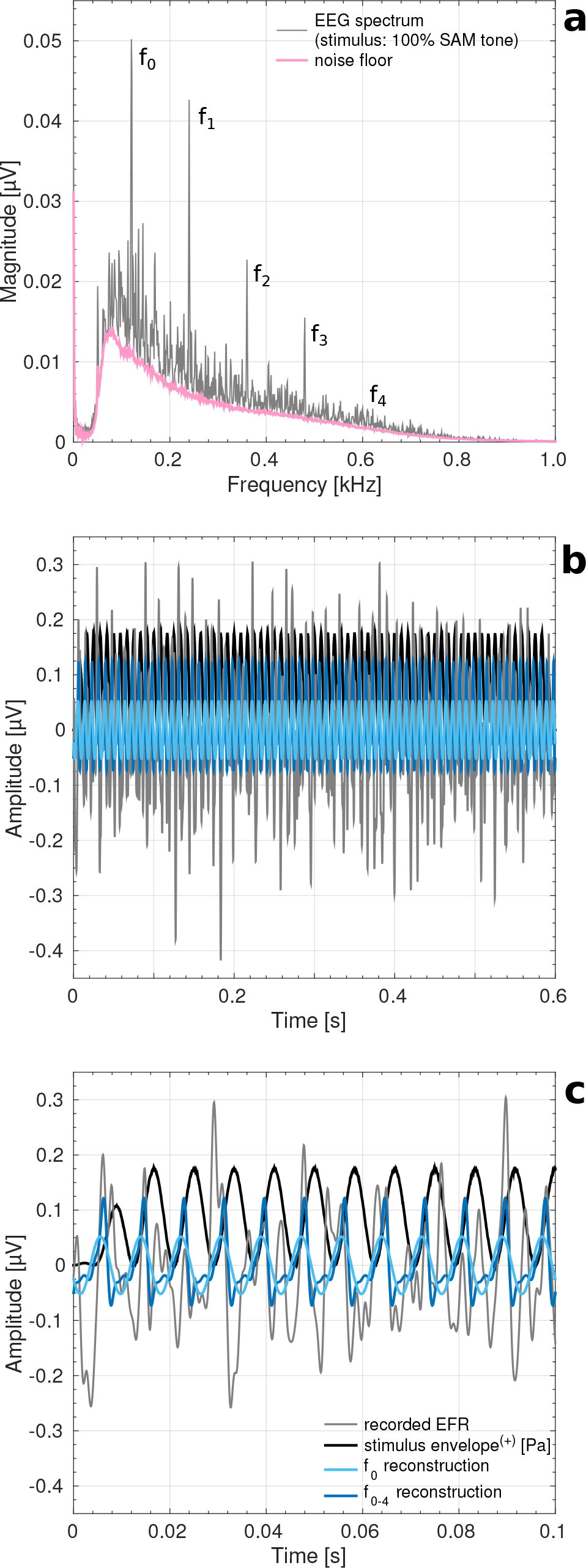
**a** Magnitude spectrum (gray) of the AEP recorded from subject NH#2 in response to a 70-dB SPL 120-Hz modulated 4-kHz pure tone. The EFR frequency response of subject NH#2 showed strong EFR components at the fundamental (f_0_) and harmonics (f_1_-f_4_) of the stimulus AM frequency. The estimated frequency dependent noise floor is depicted in pink. **b** 600 ms (i.e., the whole epoch duration) and **c** 100 ms-scaled time domain representation of the averaged EFR waveform of subject NH#2 (gray). Time-domain EFR reconstructions based on f_0_ (light-blue) and a sum of f_0_-f_4_ harmonics with the corresponding phase angle values (dark blue) are also shown. The positive part of the stimulus envelope (indicated by ^(+)^) in [Pa] is superimposed in black.

The EFR magnitude definition in Eq.1 differs from how existing studies have quantified it. Common approaches include using the magnitude of f_0_ (e.g., Kuwada et al., 1986, 2002; Dolphin and Mountain, 1992; Purcell et al., 2004; Dimitrijevic et al., 2016; Shaheen et al., 2015; Bharadwaj et al., 2015; Guest et al., 2018), the signal-to-noise ratio of the f_0_ (i.e., a relative EFR metric; van der Reijden et al., 2004; Luts et al., 2006; Garrett and Verhulst, 2019), or the phase-locking value to f_0_ (Zhu et al., 2013; Bharadwaj et al., 2015; Shaheen et al., 2015).

Figure 3b shows the benefit of using a Fourier analysis approach to reconstruct the recorded EFR waveform (gray) using Eq.1 for all available harmonics (f_0_-f_4_; dark-blue) against reconstructing the waveform from the f_0_ magnitude peak (light blue). Figure 3c shows a magnification of both recorded and reconstructed EFRs, and shows that the proposed signal reconstruction method captures the overall amplitude of the response, as well as its response peaks which are much sharper than the stimulus envelope. Aside from improved signal-reconstruction quality, the EFR_PtN_ metric in Eq.1 is a relative metric which subtracts the individual noise floor from the spectral peaks to maintain the purely stimulus-driven response. This approach minimizes how inter-individual confounds such as head size, sex, and neuro-electrical noise can affect AEP magnitudes (Trune et al., 1988; Mitchell et al., 1989) and may improve the sensitivity of the metric in quantifying individual differences in synaptopathy.

### Behavioral AM detection experiment

A subset of subjects with audiograms depicted in Fig. 2b,c also participated in a behavioral experiment which assessed their amplitude-modulation detection threshold (AM DT, i.e. the minimum AM depth for which an AM tone can be differentiated from a pure tone of the same level). Insert ER-2 headphones (Eytmotic) connected to a TDT-HB7 (Tucker-Davis) and Fireface UCX Soundcard (RME) were used for sound delivery. 500-ms long sinusoidally amplitude-modulated (100-Hz) 4-kHz pure-tones or pure-tones of 70 dB SPL were presented (with inter-stimulus intervals of 500 ms) to the same ear as tested in the AEP study. An alternative-forced choice (AFC) procedure with a 1-up-2-down protocol was followed to determine the AM DT (Levitt, 1971), and the threshold was calculated as the mean over the last six reversals at the smallest step size (i.e., 1 dB). The initial modulation depth (MD) was 50% (i.e. −6 dB re 100% modulation) and varied adaptively with step-sizes of 10, 5, 3, 1 dB across a single trial. To remove loudness cues between the reference pure-tone and AM stimuli in the AFC task, the rms levels of the reference and AM stimuli were normalized for different MDs. AM DTs were determined in one practice trial and in three trial repetitions from which the average AM DT and standard deviation was computed. It should be noted that the modulation frequency of the AM detection task was 100 Hz and not 120 Hz as used for the AEP recordings. We only realized after the start of the behavioral study that adopting a modulation frequency with a multiple of 50-Hz might be prone to line-noise artifacts in the AEP recordings. We avoided this potential confound by increasing the modulation frequency of the AEP stimuli to 120 Hz, and motivate that this difference is not expected to yield substantial differences in the interpretation of results. EFRs to 100/120-Hz SAM tones were both shown to be consistent with sources stemming from the IC or more peripheral generators based on an EFR group delay analysis (Kuwada et al., 2002; Purcell et al., 2004). Secondly, the sidebands of 100/120-Hz SAM tones were shown to fall within a single perceptual auditory filter at 4 kHz (i.e., equivalent rectangular bandwidth of 457 Hz; Glasberg and Moore (1990); resulting in unresolved components; Kohlrausch et al., 2000).

## Results

### EFR magnitude and its relationship to behavioral AM detection

We use EFR magnitudes as a marker which is sensitive to cochlear synaptopathy based on results from animal studies which related EFR magnitudes to histologically quantified synaptopathy (Shaheen et al., 2015; Parthasarathy and Kujawa, 2018), we adopt the EFR_PtN_ metric for human recordings (Eq.1) which removes the individual noise-floors and regards all available harmonics to maximize signal-to-noise ratio (Fig. 3). First, we tested whether the proposed EFR_PtN_ marker of synaptopathy maintained a relationship to behavioral temporal envelope encoding sensitivity as characterized by the amplitude-modulation detection threshold. An earlier study showed that the EFR magnitude (defined using f_0_) predicted the AM detection threshold in listeners with normal audiograms, and we repeated this experiment for a subset of listeners to assess whether our EFR marker remains predictive of a behavioral marker of temporal envelope encoding in the same listeners.

The relationship between AM DTs (in dB re 100% modulation) and different EFR metrics was investigated in Fig. 4 for f_0_-based EFR magnitudes without (panel a) or with noise-floor correction (panel b), or for EFR_PtN_ magnitudes based on Eq.1 (panel c). Where the f_0_-based EFR magnitude did not predict performance in the AM DT task (Pearson correlation R=-0.19, p=0.48, n=17), the noise-floor correction in the (f_0_-NF_0_)-based EFR magnitude improved the sensitivity of the relationship (Pearson’s R=-0.4, p=0.12, n=17). This result corroborates the initially reported relationship between the EFR (for −4 dB modulated stimulus) and the AM DT in NH listeners (Bharadwaj et al., 2015). Moreover, after applying the proposed EFR_PtN_ metric (Fig. 4c), a stronger significant correlation (Pearson’s R=-0.57, p=0.017, n=17) was observed between AM DTs and the EFR_PtN_ metric. Therefore, approximating the temporal envelope of the time-domain EFR signal by adding up the f_0_ and available harmonics can predict AM DTs in NH and HI listeners and the EFR_PtN_ can hence be considered as a relative and perceptually relevant neural marker of subcortical temporal envelope encoding (for comparable stimuli presented over the same setup).

**Figure 4.**
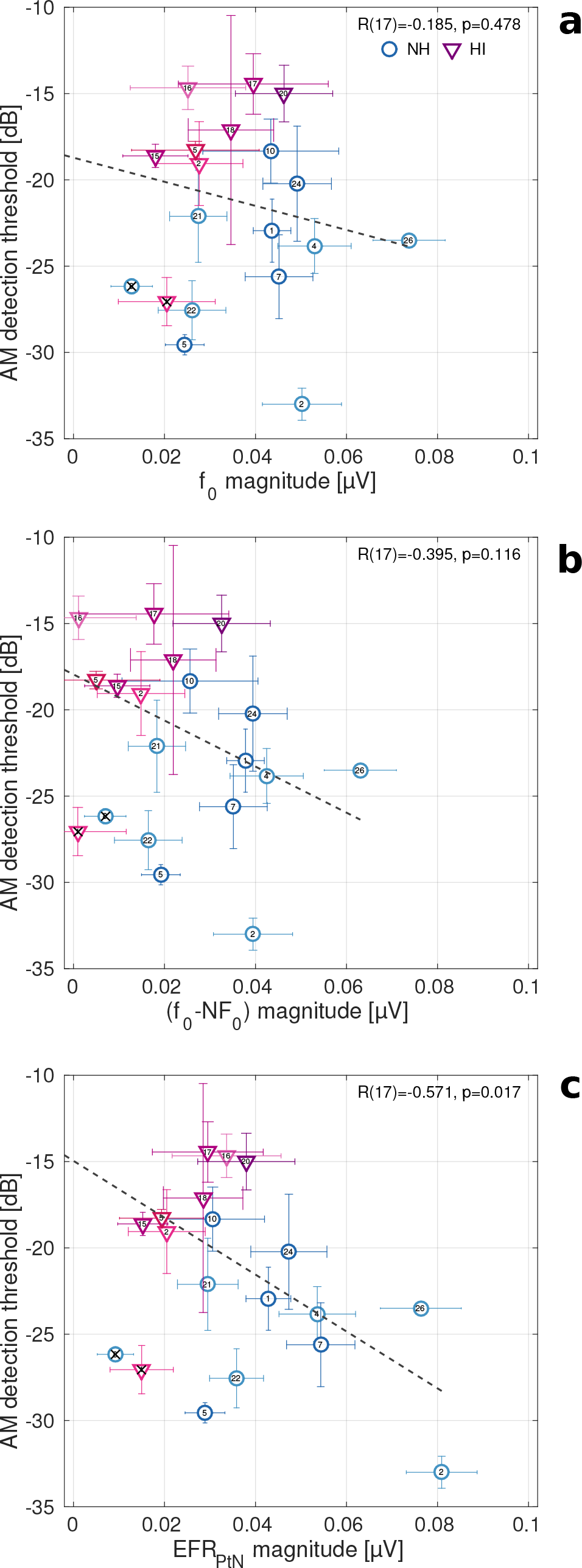
Relationship and corresponding Pearson correlation coefficient between individual AM detection thresholds (in dB re. 100% modulation) and EFR magnitudes calculated from **a**, the f_0_-magnitude of the raw EEG spectrum, **b**, the f_0_-magnitude with subtracted NF_0_ and **c**, the sum of available harmonic components at f_0_-f_4_ with the corresponding phase information and subtracted NF values (see Eq.1). The circles and triangles correspond to NH and HI subjects with the degree of audiometric threshold elevation at 4 kHz depicted (in agreement with Figure 2b,c) by the blue color gradient (from darker to lighter blue) and red color gradient (from light to dark red), respectively. Data from the two crossed symbols were omitted from the correlation analysis due to a weak EFR_PtN_ signal (after NF subtraction) for the 100% modulated SAM tone only, while stronger magnitudes for more shallow 63% and 40% conditions were seen in these subjects. This pattern of results was not observed in reference studies (Bharadwaj et al., 2015; Guest et al., 2018) and point to recording problems for these particular subjects in the 100% modulation depth condition.

### EFR sensitivity to sensorineural hearing loss

Because it is presently not clear whether degraded EFR_PtN_ magnitudes solely reflect synaptopathy or whether OHC damage also plays a role, we simulated their respective contribution to the EFR. Figures 5a,b show simulated EFR_PtN,PT_ and EFR_PtN,BB_ magnitudes to a 120-Hz SAM stimulus with a 4-kHz tone or broad-band white noise carrier, respectively, for different configurations of synaptopathy (different symbols) and cochlear gain loss (along the x-axis). The simulated cochlear gain loss profiles correspond to those in Fig. 2a and simulated EFR magnitudes for the reference NH model (no synaptopathy and no cochlear gain loss) are marked by the green filled circle in Fig.5a,b. In line with experimental data from animal synaptopathy studies (Shaheen et al., 2015; Parthasarathy and Kujawa, 2018), the simulations show that increasing the degree of synaptopathy resulted in smaller EFR magnitudes. In the model, these EFR_PtN_ reductions resulted from fewer intact IHC-AN synapses which yielded a reduced input to the CN and IC processing stages involved in the EFR generation. A loss of all simulated LSR (15% of the AN population) and MSR (15% of the AN population) fibers (HI synapt_0L,0M,13H_ profile) yielded a 20% reduction of the NH EFR_PtN,PT_ and a 22% reduction of the NH EFR_PtN,BB_. An additional loss of 50% of the HSR fibers (HI synapt_0L,0M,07H_) reduced the EFR_PtN,PT_ magnitude by an additional 37% and EFR_PtN,BB_ by 36%, showing that HSR AN fibers are more important for determining the EFR_PtN_ magnitude than LSR and MSR AN fiber types. At the same time, and in agreement with experimental studies (Kale and Heinz, 2010; Dimitrijevic et al., 2016), simulated cochlear gain loss had an opposite effect and increased the EFR_PtN,PT_ by 17% and EFR_PtN,BB_ by 45% for the simulated audiometric profiles with low-to-moderate high-frequency sloping loss. The contribution from different tonotopic locations along the cochlear partition (i.e., as characterized by the AN fibers’ CF) to the EFR is important to understand this result.

**Figure 5.**
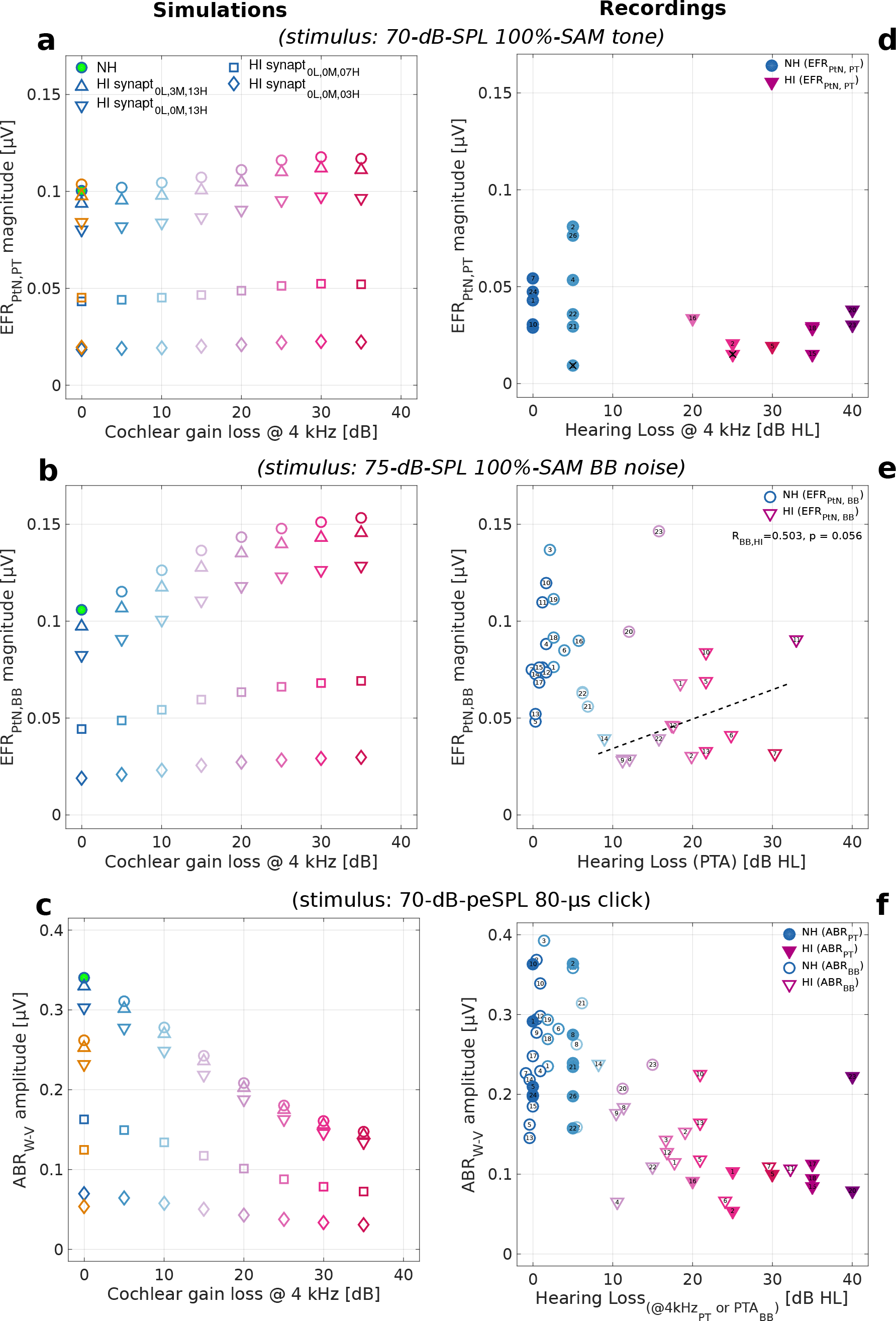
**a,d**: EFR_PT_ magnitude evoked by SAM tone, **b,e**: EFR_BB_ magnitude evoked by SAM broadband noise, and **c,f** : click-evoked ABR wave-V amplitude as a function of cochlear gain loss for the recordings (**right**) and model simulations (**left**). Open and filled symbols illustrate the AEPs as a function of threshold elevation at 4-kHz and pure tone average (PTA) across standard audiometric frequency range, respectively. Simulations were performed for different synaptopathy profiles: no synaptopathy (circles), no LSR fibers (upward triangles, HI synapt_0L,3M,13H_), no LSR&MSR fibers (downward triangles, HI synapt_0L,0M,13H_), LSR&MSR and 50% HSR fibers removed (squares, HI synapt_0L,0M,6H_), LSR&MSR and 80% of HSR fibers removed (diamonds, HI synapt_0L,0M,3H_). **Simulations**: The reference NH model (no cochlear gain loss or synaptopathy) is depicted by the green filled marker and the color gradient reflects cochlear gain loss degree (abscissa values). **Recordings**: circles and downward triangles indicate EFR_PtN_ magnitudes for NH and HI subjects, respectively. Recordings from two subjects depicted with crossed symbols and near-zero EFR_PtN_ were excluded from the ABR_W-V_/EFR_PT_ ratio calculation as well as SNHL map construction in Fig. 8d,e.

Figure 6 illustrates how different CFs of simulated cochlear excitation patterns to the PT or BB SAM stimulus (panel a) contribute to AM encoding in the IC (panel b) for simulated normal-hearing (dark-blue) or high-frequency cochlear gain loss (dark-red) and different synaptopathy profiles (different symbols). In agreement with several studies (Encina-Llamas et al., 2019; Shaheen et al., 2015; Joris and Yin, 1992), the model predicts that the relative contribution of off-CF fibers (approximately with CFs*>* 6 kHz for the 70-dB PT stimulus) is greater than that of on-CF fibers for the NH EFR. On-CF HSR fibers have the largest input (see Fig. 6a) and hence operate close to the saturated region of the AN rate-level function (Liberman, 1978; Joris and Yin, 1992). Differently, off-CF HSR fibers receive attenuated cochlear excitation input and operate in the non-saturated region of the rate-level curve, which causes HSR fibers to capture AM information in the low-level tails of the cochlear excitation pattern. Thus, even though LSR and MSR fibers contribute considerably to on-CF AM sensitivity, their contribution to the overall EFR_PtN,PT_ magnitude (which reflects AM information across CF) was only in the order of 20% (Fig. 5a, synapt_0L,0M,13H_). Due to the prominent contribution of HSR fibers to the EFR magnitude, removing a 50% of available HSR fibers (synapt_0L,0M,7H_) reduced the EFR magnitude strongly and, resulted in halving the simulated EFR_PtN,PT_ (Fig. 5a).

**Figure 6.**
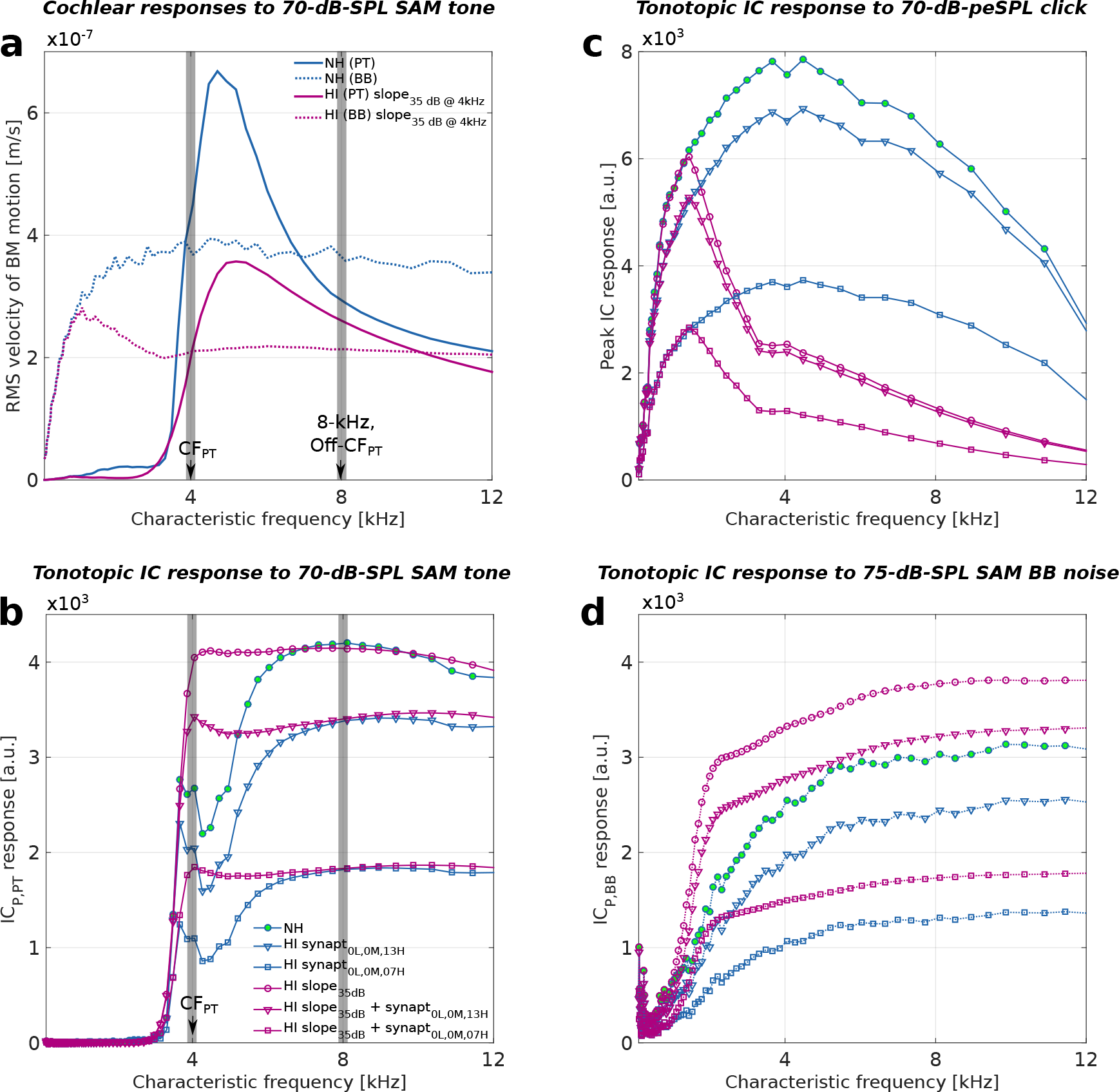
**a** Simulated BM motion; **b,d** modulated and **c** peak IC responses across CFs evoked by a 70-dB SPL 120-Hz SAM tone with a 4-kHz carrier (PT), a 75-dB SPL 120-Hz SAM broadband white noise (BB) and a 70-dB-peSPL, 80*µ*s click, respectively. Simulations were performed for the normal-hearing model (NH; dark-blue), high-frequency sloping cochlear gain loss (HI; dark-red) as well as different degrees of synaptopathy (HI; different symbols).

Simulated cochlear gain loss mostly increased AM contributions from the on-CF region and yielded a greater relative contribution of on-CF versus off-CF fibers than observed for the NH profile (Fig. 6b,d; compare red and blue PT traces). The enhanced on-CF AM encoding resulted from a reduced and linearized cochlear input to the IHC/AN complex which caused AN fibers to operate in a more sensitive range for AM encoding at fixed stimulation levels (see Fig. 6c,d; and experimental observations in Joris and Yin, 1992; Kale and Heinz, 2010). The BB SAM excitation pattern was broader than the PT SAM pattern and hence cochlear gain loss resulted in stronger EFR enhancement for the BB than PT carrier (compare Fig. 5b and Fig. 5a). Consequently, for the same degree of cochlear synaptopathy, Fig. 5a,b shows that both simulated EFR_PtN,PT_ and EFR_PtN,BB_ magnitudes increased when greater degrees of high-frequency cochlear gain loss were introduced. The EFR enhancement associated with cochlear gain loss became smaller for larger degrees of synaptopathy (Fig. 5a,b; compare circles and diamonds) due to the overall smaller number of AN fibers contributing to the EFR (Fig. 6b,d). Therefore, cochlear gain loss can equalize EFR_PtN_ magnitudes towards NH values when a combination of impairments is present (e.g., compare the green circle with red upward triangles in Fig. 5a or red downward triangles in Fig. 5b). The observed equalization is particularly important for quantifying selective LSR/MSR fiber synaptopathy, whose effect on the EFR magnitude can be fully compensated by the co-existing OHC dysfunction.

Human reference EFR_PtN,PT_ and EFR_PtN,BB_ magnitudes are plotted as a function of the audiometric hearing threshold in Fig. 5d,e for listeners with normal (blue gradient) and hearing-impaired (red gradient) audiograms. For the EFR_PtN,PT_ recordings (Fig. 5d, filled symbols), the hearing threshold at 4 kHz was reported, which corresponds to the carrier frequency of the SAM tone, whereas for the EFR_PtN,BB_ results, the audiometric threshold average across the standard frequency range (PTA; see also Table 1.) was reported (Fig. 5e, open symbols). Reference EFR magnitudes were generally smaller for older HI (triangles) than for young NH (circles) participants for both stimulus paradigms: a two-sample T-test (p=0.022, passing the Anderson-Darling test for the normal-distribution assumption) was applied to test the EFR_PtN,PT_ samples and a two-sided Wilcoxon rank test (p=0.0001) was used to test the EFR_PtN,BB_ samples. At the same time, we did not observe a consistent EFR decrement as the PTA increased from 10 to 40 dB HL for HI listeners (open triangles). Instead, we observed increased EFR magnitudes which approached NH values in line with our model predictions and with AN (Kale and Heinz, 2010) and EFR (Dimitrijevic et al., 2016) studies showing increased envelope coding for subjects with elevated ABR or audiometric hearing thresholds, respectively. A correlation analysis showed a positive non-significant trend (Fig. 5e; R_BB,HI_=0.503, p=0.056, N=15) for recorded HI EFR_PtN,BB_ (for which the model predicted stronger OHC-loss-induced magnitude enhancement; Fig. 5b). Lastly, it should be noted that NH listeners with elevated audiometric thresholds (see PTA *>* 10; Fig. 5e, circles) also had strong EFR magnitudes which can degrade with age for similar PTA values (e.g. compare NH subject #20 and NH #23 with the most elevated PTA; Fig. 5e and different age; Table 1.)

### ABR sensitivity to sensorineural hearing loss

The ABR wave-I (1-2 ms after transient sound onset) is generated near the VIII^*−th*^ nerve (Picton, 2010) and its strength depends on the available AN fiber population (Kujawa and Liberman, 2009). However, wave-I is small in humans and might yield unreliable measurements for the vertex electrode configuration (Plack et al., 2016; Mehraei et al., 2016; Garrett and Verhulst, 2019). For this reason, we focus on how sensorineural hearing loss affects the ABR wave-V, which can be recorded robustly in NH and HI humans (Gorga et al., 1985; Verhulst et al., 2016; Madsen et al., 2018; Garrett and Verhulst, 2019). The ABR wave-V for NH listeners occurs 5-7 ms after sound onset and reflects the ascending input activity to the IC (Møller and Burgess, 1986; Melcher and Kiang, 1996; Picton, 2010; Bidelman, 2015). By focusing on the ABR wave-V, we thus assume that any peripheral hearing impairment affecting the Wave-I would propagate along the ascending auditory pathways and influence the ABR wave-V as well. Fig. 5c depicts simulated W-V amplitudes to a 70-dB-peSPL click for the considered hearing loss profiles (Fig. 2a) and shows that both cochlear gain loss and synaptopathy can cause ABR amplitude reductions. This result corroborates several synaptopathy studies which report degraded supra-threshold compound action potential and ABR wave-I amplitudes (Kujawa and Liberman, 2009; Furman et al., 2013; Sergeyenko et al., 2013; Bourien et al., 2014; Kujawa and Liberman, 2015; Valero et al., 2017). Additionally, selective OHC damage induced by styrene exposure was shown to reduce the compound action potential amplitude for stimulation levels below 90 dB SPL (Chen et al., 2008), in line with our reduced ABR wave-V amplitudes for cochlear gain loss. The simulations in Fig. 5b clearly show the cumulative and degrading effect synaptopathy (*≈*80% reduction for simulated synaptopathy profiles) and cochlear gain loss (*≈*55% reduction for simulated audiograms) have on the ABR amplitude. Although we consider wave-V amplitudes in Fig.5c, reductions of the same order (*≈*80% and *≈*50% for simulated synaptopathy and cochlear gain loss profiles) were also observed for the simulated ABR wave-I amplitudes (Fig.7a), and the results of this study can hence be translated to recorded ABR wave-I amplitudes of sufficient signal quality.

**Figure 7.**
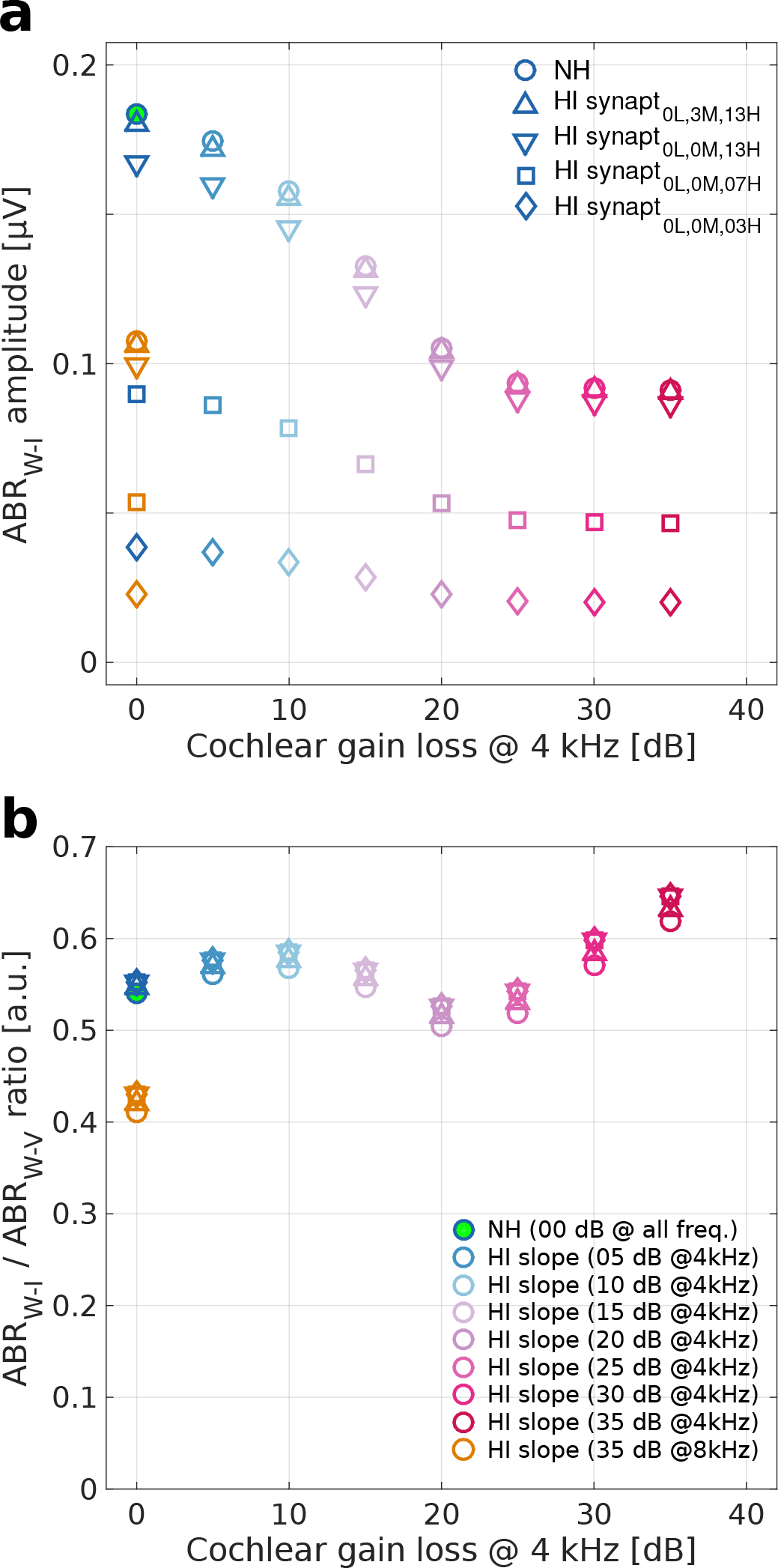
Simulated **a** ABR wave-I and **b** ABR wave-I / ABR wave-V ratio as a function of cochlear gain loss for different synaptopathy profiles.

To better understand how SNHL affects the ABR generators, Fig. 6c shows how simulated IC peak activity in different CF units contribute to the ABR. LSR/MSR fiber loss (i.e., HI synapt_0L,0M,13H_) mostly affected those CF regions which contributed strongly to the ABR in the NH model. A loss of HSR fibers (i.e., HI synapt_0L,0M,07H_) resulted in a substantial across-CF reduction in the simulated IC response, as earlier observed for the simulated EFRs. Cochlear gain loss reduced the contribution from all fiber types in the CF regions affected by the loss (Fig. 6c). ABR amplitudes can be reduced similarly by synaptopathy or OHC damage (Fig. 5c), but Fig. 6c shows that different SNHL aspects affect different CF regions. Simulated uniform across-CF synaptopathy mostly reduced the 2-5 kHz frequency contribution to the ABR because of the generally-larger IC response in this range due to the middle-ear filter properties. Differently, simulated high-frequency cochlear gain loss had a strong effect on reducing the high-frequency CF contributions.

Recorded ABR wave-V amplitudes to a 70-dB peSPL condensation click are plotted as a function of the audiometric hearing threshold in Fig. 5f: the 4 kHz threshold was used for ABRs recorded in the same session as the EFR_PtN,PT_ (Fig. 5d, filled symbols) and the pure-tone average was used for ABRs recorded in the EFR_PtN,BB_ session (open symbols). ABR W-V amplitudes were significantly greater for NH subjects than for HI subjects (open symbols: two-sample T-test, p=0.000001, passing the Anderson-Darling test for normality; filled symbols: two-sided Wilcoxon rank test, p=0.0008), and fell in range with simulated amplitudes for the different SNHL profiles.

### Isolating cochlear gain loss from evoked potentials

Because the simulations in Fig. 5a,b showed that both cochlear gain loss and synaptopathy affect simulated ABR and EFRs, another approach is required to adopt AEPs as a sensitive diagnostic tool for synaptopathy when potential OHC damage is also present. Specifically, if EFR magnitudes can be considered together with a metric which is only sensitive to OHC damage in the individual listener, the synaptopathy and OHC aspects of SNHL can be separated. To this end, we first introduce an AEP ratio which is sensitive to supra-threshold OHC impairment and insensitive to synaptopathy:

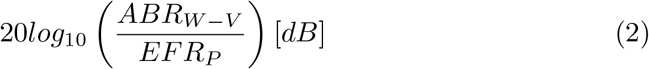

No noise-floor correction was applied to yield the EFR_P_ in the ratio, as the individual noise floor is expected to contribute similarly to both the EFR and ABR metrics in the same recording session. The proposed ratio and its sensitivity to OHC damage is based on the following arguments: (i) Synaptopathy was in Figs. 5a,b shown to reduce the EFR and ABR amplitudes similarly, which means that its influence will not affect the ratio. As can be observed from the simulations in Fig. 8 (a,b,c), synaptopathy only has a minor influence on the ratio and the observed ratio decrements are mainly caused by cochlear gain loss. (ii) any non-hearing-related individual characteristics contributing to both the EFR and ABR (e.g., sex, head size, noise floor) are canceled in the ratio. Lastly, (iii), any compensatory gain mechanism which could yield normal wave-V amplitudes for reduced wave-I amplitudes after AN fiber reduction (e.g., homeostatic gain changes; Schaette and McAlpine, 2011; Chambers et al., 2016; Möhrle et al., 2016), would affect both the EFR and ABR wave-V generators which stem from similar stages of the auditory system (Melcher and Kiang, 1996; Picton, 2010; Bidelman, 2015) and would hence cancel out in the ratio.

**Figure 8.**
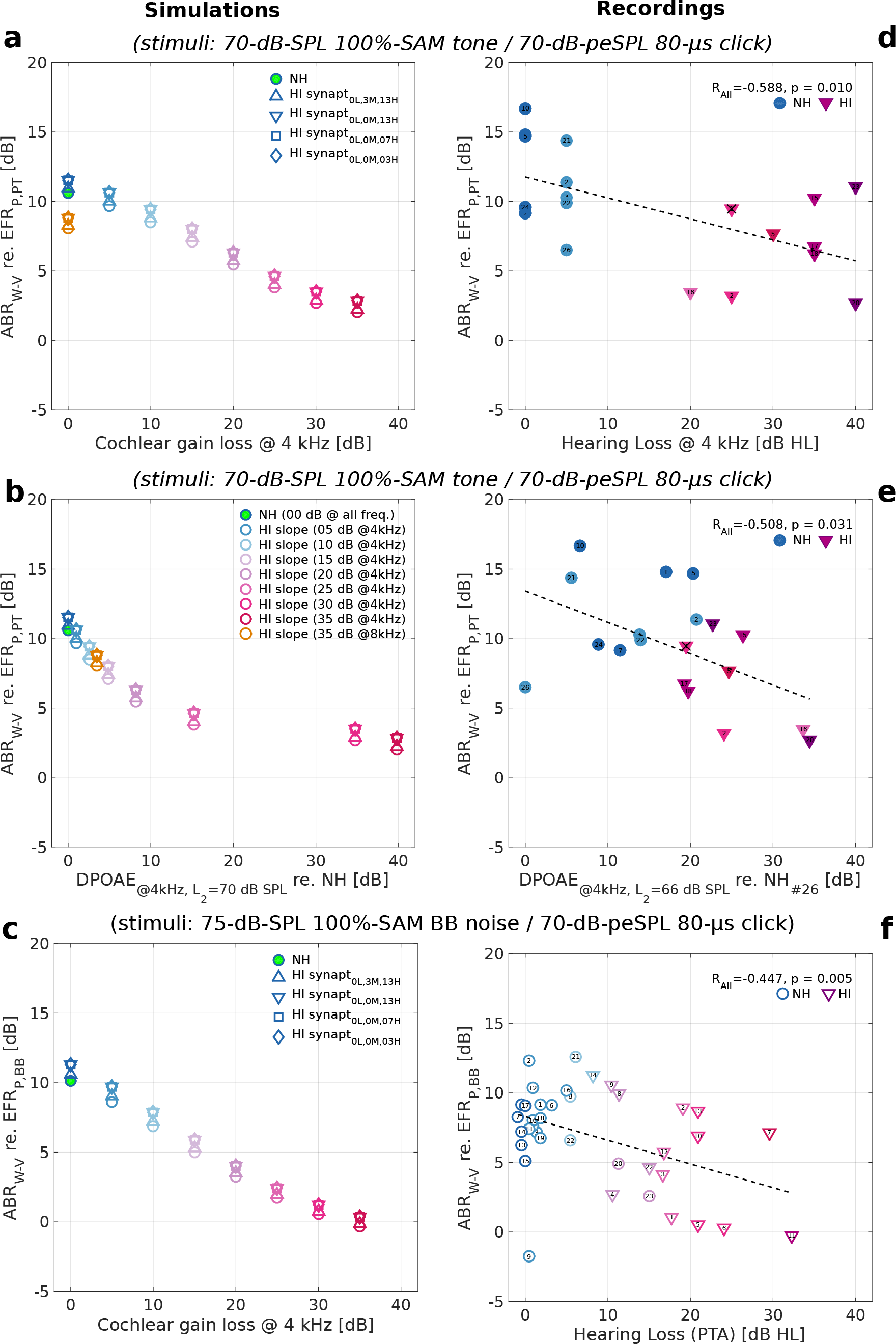
Simulated (**left**) and recorded (**right**) ABR_W-V_/EFR_P_ ratios as a function of **a,d** cochlear gain loss at 4-kHz; **b,e** supra-threshold DPOAE magnitude shifts at 4-kHz (re. model NH profile, L_2_ = 70 dB SPL or the best measured NH DPOAE, L_2_ = 66 dB SPL); and **c,f** pure tone average (PTA) across standard audiometric frequency range.

The cochlear gain loss degree estimated using the ABR_W-V_/EFR_P_ ratio refers to *supra-threshold* gain loss associated with OHC damage and targets similar cochlear frequency regions as the adopted supra-threshold EFR_PtN_ metric for synaptopathy. The ratio estimates gain loss using a 70-75 dB stimulus, hence the estimated OHC loss is not necessarily identical to the loss estimated using DPOAE, ABR, or behavioral hearing threshold metrics. While OHC damage can reduce BM vibration across the entire stimulus level range, BM response reductions are different at low and high stimulus intensities (e.g. see input-output function slope; Ruggero and Rich, 1991). It is also expected that the ABR_W-V_/EFR_P_ ratio is sensitive to the shape of the audiogram due to the broad tonotopic range which contributes to AEPs for high SPLs. The simulations in Fig.8a illustrate this point by showing that steeper high-frequency cochlear gain loss slopes can yield different ABR W-V/EFR ratios even though the audiometric thresholds at 4-kHz (corresponding to the EFR_PT_ stimulus) were matched (e.g. 0 dB at 4 kHz and 35 dB at 8 kHz; compare dashed orange vs solid dark blue).

From the supra-threshold gain perspective, we further explored the theoretical relationship (Fig.8b) between the ABR_W-V_/EFR_P_ ratio and the simulated supra-threshold DPOAE magnitude shift (L_2_ = 70 dB SPL corresponding to the SAM PT stimulus intensity) for different cochlear gain loss configurations and compared our simulations against recorded data (Fig.8e, L_2_ = 66 dB, i.e. max L_2_ for both NH and HI participants). As predicted, smaller ratios were obtained for larger degrees of sloping cochlear gain loss, although the relationship was not linear for the model (Fig.8b). Applying the ABR_W-V_/EFR_P,PT_ ratio to the study participants (Fig. 8e) yielded a negative trend and, overall, the experimental values were in range with the simulated ABR/EFR ratio and supra-threshold DPOAE magnitudes.

Lastly, to better match the cochlear frequency regions targeted by the ABR and EFR stimuli in the ratio, we also calculated the ABR/EFR ratio from the EFR_P,BB_ which was evoked by SAM broadband noise and had a tonotopic contribution comparable to that evoked by the click (compare Fig.6c and Fig.6d). In agreement with the theoretical prediction, both simulated (Fig.8c) and recorded (Fig.8f) ABR_W-V_/EFR_P,BB_ ratios decreased as audiometric threshold increased, showing a significant negative trend for the human data. Low-frequency audiometric loss did not substantially influence the EFRs and ABRs for the considered stimulus parameters (see reduced tonotopic contribution for low CFs; Fig.6c,d), while the PTA metric did capture these low-frequency hearing sensitivity differences. This mismatch between the PTA and estimated OHC loss from AEPs might explain the observed variability across ABR_W-V_/EFR_P,BB_ ratios for HI listeners with similar PTAs (e.g. compare ratios for HI #2, 10, 13 and for HI #1, 5 with the similar PTA *≈* 20 dB, but elevated or normal low-frequency audiometric thresholds, respectively; Fig.8f and see Table I for across-frequency audiometric thresholds).

### A Sensorineural Hearing Loss Map (SNHL Map)

Even though the largest EFR_PtN_ magnitude decrements were associated with synaptopathy in the model simulations (Fig. 5a,b), cochlear gain loss was also seen to affect this metric and can compensate the EFR magnitude decrement caused by cochlear synaptopathy as the degree of OHC damage increases. Consequently, we propose to use the EFR in combination with a second AEP metric which is sensitive to cochlear gain loss (i.e., the ABR_W-V_/EFR_P_ ratio) to yield a correct interpretation of the supra-threshold EFR_PtN_ marker in terms of synaptopathy.

Figure 9 shows that combining the EFR_PtN_ marker with the ABR_W-V_/EFR_P_ratio (or supra-threshold DPOAE magnitude) can separate the simulated hearing pathologies, where deviations along the horizontal dimension capture variations in the synaptopathy degree and where deviations along the vertical dimension capture individual differences in supra-threshold cochlear gain loss. The normal-hearing reference (circles in the model-derived scheme; Fig. 9d) is characterized by: (i) a strong EFR_PtN_ magnitude which is smaller than for cochlear gain loss profiles without synaptopathy, and (ii), a high DPOAE magnitude (Fig. 9c) or large ABR_W-V_/EFR_P_ ratio which is somewhat smaller than for selective LSR/MSR synaptopathy. The latter fiber types contribute more strongly to the sustained EFR response than to the ABR_W-V_ resulting in an imperfect cancellation of synaptopathy in the ratio.

**Figure 9.**
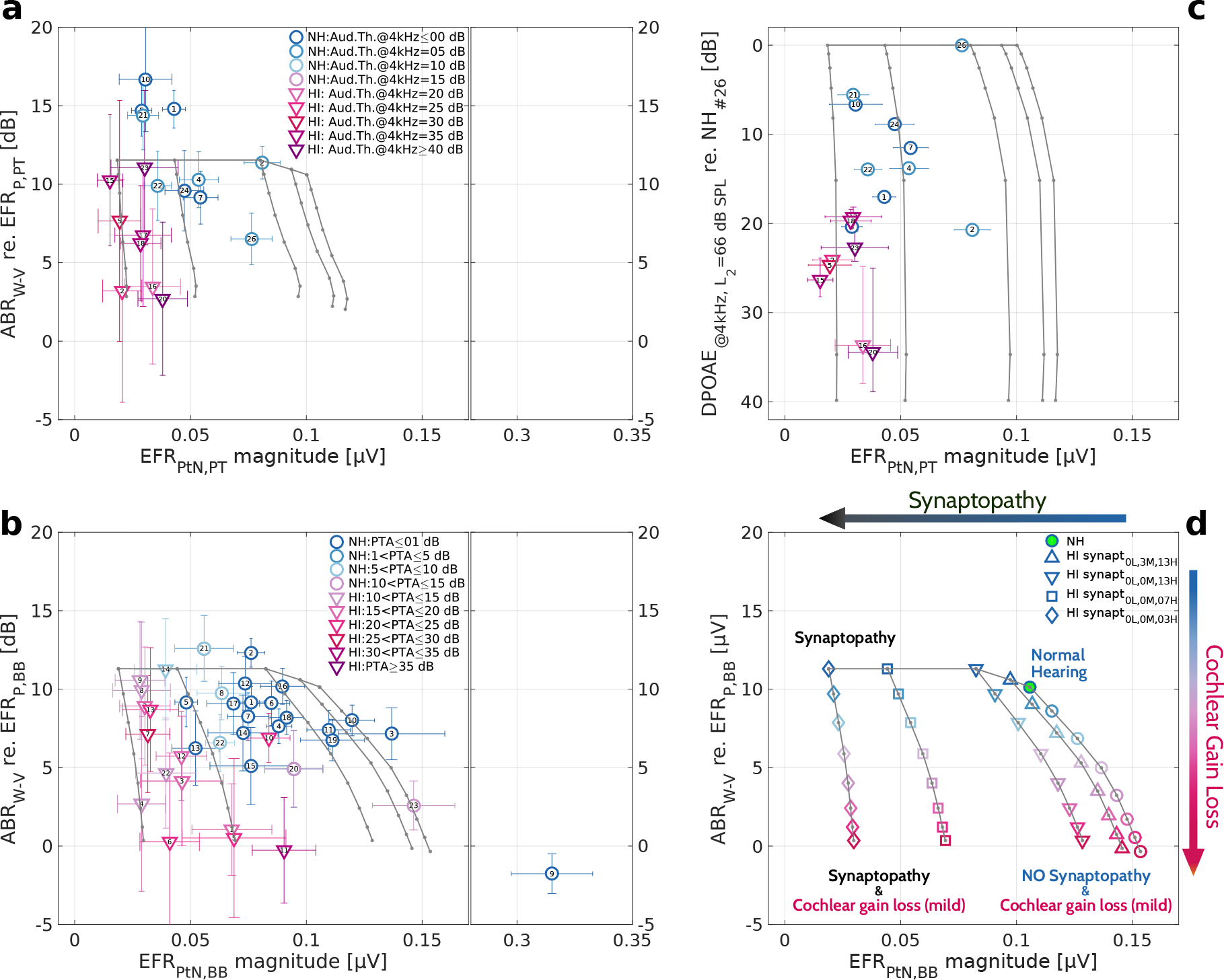
Simulated (overlaid grids) and reference sensorineural hearing loss maps based on **a,b** the ABR_W-V_/EFR_P_ ratio or **c** supra-threshold DPOAE magnitude shifts at 4-kHz and EFR_PtN_ magnitude evoked by **a,c** 70-dB SPL 120-Hz SAM tone with a 4-kHz carrier or **b** 75-dB SPL 120-Hz SAM broadband noise. **d** Colored scheme based on supra-threshold BB stimuli which summarises four main extremes on the SNHL map: the NH reference and combinations of synaptopathy and cochlear gain loss associated with OHC damage.

Based on the model simulations in Fig. 9 (see overlaid model-derived grids in a-c), it is possible to map an individual on the SNHL map as follows: (i) determine their vertical position according to the OHC-sensitive ABR_W-V_/EFR_P_ ratio, and (ii), determine their horizontal direction based on the EFR_PtN_ magnitude. The closest vertical gridline to the mapped data-point will yield the individual synaptopathy profile. Figure 9a shows a SNHL map for subjects with audiograms in Fig. 2b,c using the same stimulus protocols as adopted for the model simulations: i.e., a 70-dB-peSPL, 80-*µ*s click and a 70-dB-SPL 120-Hz 100%-SAM tone with 4-kHz carrier. The color scheme illustrates different degrees of cochlear gain loss at 4 kHz according to the color scheme adopted in Figs. 2b,c. There is qualitative agreement between the model simulations (see model-derived grid) and reference data, however, individual absolute EFR_PtN,PT_ magnitudes were overall smaller than simulated. Nevertheless, the general trends of how the experimental metrics changed when SNHL was introduced, corroborated the predictions. Reduced ABR_W-V_/EFR_P,PT_ ratios were observed for HI compared to NH listeners, consistent with reduced hearing sensitivity associated with OHC damage. Whereas the cochlear gain loss and synaptopathy simulations yielded individual EFR_PtN_ magnitudes spanning the whole horizontal range of recorded EFR_PtN_ magnitudes (i.e. from no to severe synaptopathy), the HI reference data clustered near the bottom left of the SNHL map. This clustering can theoretically occur when older listeners with more severe degrees of OHC loss suffer from the worst degrees of synaptopathy as well. Whereas this statement cannot be proven using the human dataset at hand, there are animal physiology studies which agree with this predictions by showing that synaptopathy occurs earlier in time than OHC damage as a consequence of the aging process (Sergeyenko et al., 2013; Fernandez et al., 2015; Parthasarathy and Kujawa, 2018).

We also constructed a SNHL map with simulated and recorded DPOAEs for the same listeners as in Fig. 9a to compare SNHL maps constructed using supra-threshold DPOAE (Fig. 9a) or AEP metrics (Fig. 9a). Consistent with the observed ABR_W-V_/EFR_P,PT_ ratio reduction, Fig. 9c shows degraded supra-threshold DPOAE magnitudes for HI listeners with elevated audiometric thresholds.

To optimize the accuracy of the ABR_W-V_/EFR_P,PT_ metric, it is crucial that both metrics in the ratio have comparable cochlear sources (and CFs) contributing to the population response. This was not the case for the EFR_P,PT_ evoked by a 4-kHz SAM tone used in combination with the broadband click ABR which had generators from a broad CF range down to 0.5-1 kHz for suprathreshold stimulation (Fig. 6c; Eggermont and Don, 1980; Abdala and Folsom, 1995). To account for this potential incorrect cancellation, we constructed a SNHL map in which an EFR_P,BB_ metric was derived from the response to a 75-dB-SPL 100%-SAM broadband white noise (Fig. 9b). This stimulus yielded a broad tonotopic excitation along the BM similar which was similar to that of a click stimulus (compare Fig. 6c and d) Simulated and recorded EFR_BB_ magnitudes were overall larger compared to the EFR_PT_ response and off-CF cochlear regions contributed less to the potential mismatch between simulated and reference EFRs (compared to the EFR_PT_) yielding comparable absolute EFRs_BB_ magnitudes (Fig.5b and e). The simulated SNHL map for the EFR_BB_ metric (Fig. 9b(grid),d) demonstrated similar trends to the SNHL map constructed for the EFR_PT_ (Fig. 9a(grid)), while preserving a good quantitative agreement with the reference data. Figure. 9b shows a human SNHL map using the EFR_P,BB_ metric for a larger group of study participants (n=38, 23 NH and 15 HI listeners; Table I) than which participated in the EFR_P,PT_ experiment. The simulated degree of synaptopathy (from NH to HI synapt_0L,0M,03H_) captured the horizontal spread in individual EFR_PtN,BB_ magnitudes well and individual ABR_W-V_/EFR_P,BB_ ratios were clustered within the predicted range for the simulated sloping high-frequency hearing loss profiles. Simulated EFR_BB_ magnitudes showed a notable rightward skewing as cochlear gain loss increased for the NH or synaptopathy models (Fig. 9b(grid),d). Consequently, the EFR_PtN,BB_ magnitude can be greater for increasing cochlear gain loss degrees and compensate co-existing degrees of synaptopathy (e.g. compare green-filled symbol and dark-red downward triangles; Fig. 9d). This effect is caused by the interplay between (i) OHC damage increasing the EFR_PtN,BB_ as a consequence of gradually more unsaturated AN fibers, and (ii), by removing those saturated AN fibers as a consequence of increasing degrees of synaptopathy (compare lines of the same color with different markers in Fig. 6d). In agreement with the simulated rightward skewing, the NH and HI data showed enhanced EFR_PtN,BB_ magnitudes as the ABR_W-V_/EFR_P,BB_ ratio decreased. Moreover, similar than for the EFR_PT_-based SNHL map, the HI data clustered towards the bottom left corner of the SNHL map indicating co-existing synaptopathy and OHC damage. To summarize, two independent measurement sets showed the same trend in both SNHL maps and captured how the SNHL map is predicted to alter when the stimulus characteristics changed from PT to BB.

Figure 9d schematically summarizes four main extremes on the SNHL map with supra-threshold AEP metrics based on the EFR_BB_ condition. The horizontal direction reflects cochlear synaptopathy changes while the vertical direction estimates the degree of supra-threshold cochlear gain loss. The exact shape of the map is influenced by the bandwidth and level of the adopted EFR and ABR stimuli (e.g., see difference between EFR_PtN,PT_ and EFR_PtN,BB_ maps; Fig. 9a,b, grids). The extremes of the SNHL map reflect the following four categories: *Normal Hearing* : NH participants show large ABR_W-V_/EFR_P_ ratios and high EFR_PtN_ magnitudes which cluster towards the top-right corner of the SNHL map (green filled marker). *Synaptopathy* : participants with synaptopathy but no OHC dysfunction show reduced EFR_PtN_ magnitudes and slightly larger than normal ABR_W-V_/EFR_P_ ratios which cluster towards the top-left corner of the map. *No Synaptopathy & Cochlear gain loss* results in reduced ABR_W-V_/EFR_P_ ratios and enhanced EFR_PtN_ magnitudes due to OHC damage in the absence (or for a fixed degree) of synaptopathy (SNHL profiles cluster towards the bottom-right of the map). *Synaptopathy & Cochlear gain loss*: In the presence of OHC damage, the degree of synaptopathy can be established by first mapping the ABR_W-V_/EFR_P_ ratio in the vertical direction, after which the corresponding EFR_PtN_ magnitude determines the degree of synaptopathy.

## Discussion

### Sensitivity of the SNHL map to different stimulus characteristics

From a sensitivity point of view, it is possible to further optimize the stimulus characteristics and yield AEP metrics in the ABR_W-V_/EFR_P_ ratio stemming from identical cochlear regions e.g. by combining the EFR_PT_ with tone-burst ABRs. The SNHL maps presented here offer a single quantification of supra-threshold cochlear gain loss and synaptopathy across a rather broad tonotopic region, whereas it might be possible to build a range of frequency-specific SNHL maps using bandwidth-limited stimuli. EFR magnitudes to 100% modulated stimuli were shown to be an effective metric to build the SNHL map, but the concept could be further refined to target sensitivity to different subtypes of synaptopathy (e.g. by using the EFR slope metric targeting LSR fibers; Bharadwaj et al., 2014, 2015). Given that a considerable amount of off-CF HSR fibers were shown to contribute to the supra-threshold SAM-tone evoked EFR_PtN,PT_ (Fig. 6b; Encina-Llamas et al., 2019), presenting the stimuli in different types of noise maskers might also be an option to suppress off-CF HSR contributions and yield an EFR metric which reflects the on-CF fiber contributions more strongly (Bharadwaj et al., 2014). Whether more advanced band-limited SNHL maps (using e.g., tone-bursts, narrow-band noise carriers, masking noise) would yield satisfactory evoked potential signal quality in NH and HI human listeners remains to be explored.

### Potential central gain compensation

We propose the use of the ABR wave-V in our ABR_W-V_/EFR_P_ ratio, but several studies have suggested that homeostatic gain can increase along the ascending auditory pathway after synaptopathy or IHC lesions (Schaette and McAlpine, 2011; Möhrle et al., 2016; Chambers et al., 2016; Hesse et al., 2016) and yield normal wave-V amplitudes in the presence of reduced wave-I amplitudes. The central gain hypothesis questions the use of ABR waves beyond wave-I for synaptopathy diagnosis. However, the data on homeostatic gain is still sparse and several studies report evidence that synaptopathy can reduce both the supra-threshold ABR wave-I and later wave-V/(IV) amplitudes. Even though the reductions were less strong for the wave-IV, than wave-I the Hesse et al. (2016) study shows that *both* the supra-threshold IC firing rate (strongly associated with the generation of the ABR wave-V(IV) complex; Picton, 2010) and the ABR wave-IV were reduced compared to control recordings after noise-induced AN deafferentation without permanent hearing-threshold shifts. Similarly, the Chambers et al. (2016) ouabain-induced cochlear denervation study shows that IC firing rates did not recover back to the control condition after 30 days, whereas auditory cortical responses did. From a physiological perspective, a straightforward interpretation of these findings with respect to homeostatic gain is difficult, because if brainstem/midbrain gain changes after cochlear denervation, the IC firing rate should be restored back to normal.

Moreover, central gain changes have experimentally been characterized using the ABR wave-I/V(IV) amplitude ratio which can be influenced by (i) the frequency-dependence of the ABR generators (i.e. the CF mismatch between wave-I and wave-V(IV) generators; Don and Eggermont, 1978), and (ii), the frequency-specific audiogram shape (Verhulst et al., 2016). This means that a direct interpretation of the ABR wave-I/V(IV) ratio might be difficult without a careful estimation of the audiometric thresholds (including EHFs) for NH subjects and older listeners with elevated hearing tresholds. The model we adopted captures the frequency-dependence of the ABR wave-I and V genera-tors (Verhulst et al., 2015, 2018) and hence predicts that OHC damage can yield ABR wave-I/V(IV) deviations, even in the absence of simulated synaptopathy or homeostatic gain. Figure 7b illustrates how the ABR wave-I/V(IV) ratio first increases and later decreases as the degree of high-frequency sloping hearing loss is increased. Additionally, there is a large difference between the ABR wave-I/V(IV) ratio for a model with a normal 4-kHz threshold which either had EHF cochlear gain loss or not (Fig. 7b; compare green-filled and yellow markers). These simulations illustrate how EHF OHC damage can influence the ratio using a model which simulated CN and IC neuron transfer functions (Nelson and Carney, 2004) but did not include specific homeostatic gain mechanisms between the AN and IC model stages (e.g., unlike the model of Schaette and McAlpine, 2011).

Based on the above arguments, and in the absence of a full physiological understanding of homeostatic gain processes and their effect on ABR waves, we refrained from introducing this concept in our model simulations. We also opted to use SNHL maps on the basis of the ABR wave-V because this response can be recorded reliably in NH and HI human listeners with higher signal-to-noise ratio than the ABR wave-I in the vertex electrode configuration (e.g., Gorga et al., 1985; Madsen et al., 2018; Garrett and Verhulst, 2019). Even if future physiology studies confirm a compensating brainstem or midbrain gain mechanism after synaptopathy, its potential effect is expected to be minimal for our SNHL maps as we consider the ABR wave-V amplitude in a ratio with the EFR_P_ magnitude. Because both responses are generated within similar subcortical stages of the auditory system (Moller, 2007; Picton, 2010; Bidelman, 2015), homeostatic gain should influence both metrics and hence cancel out in the ratio. In case homeostatic gain would only affect the ABR but not EFR_P_, the proposed ABR_W-V_/EFR_P_ ratio could be re-designed to include ABR wave-I amplitudes. This would imply adopting a more sensitive electrode configuration than conventional ABR setups provide (e.g. using ear-canal tiptrodes; Bauch and Olsen, 1990; Liberman et al., 2016; Mehraei et al., 2016; Prendergast et al., 2018)

Lastly, it might be that homeostatic plasticity restores both the mean *and* modulated firing rate of brainstem/midbrain neurons due an imbalance of excitatory and inhibitory synapses and/or intrinsic neuronal excitability (Schaette and McAlpine, 2011). In this case, homeostatic gain could also affect the synaptopathy-sensitive EFR_PtN_ metric. However, this suggestion disagrees with well-established experimental evidence showing that the modulated response decreases when the mean response rate increases for supra-threshold stimulation (e.g., Joris and Yin, 1992, see single-unit rate-level curves at CF). At the same time, several experimental studies reported reduced EFR magnitudes associated with histologically-verified synaptopathy (Shaheen et al., 2015; Parthasarathy and Kujawa, 2018) and question a possible EFR restoration mechanism related to brainstem/midbrain homeostatic gain.

### Limitations of the model-based approach

Despite the advantages of model-based approaches in exploring how various SNHL profiles affect the sensitivity of hearing diagnostic metrics within a broad parameter space, models only constitute a compromised description of the different stages and nonlinear features of the auditory system. The model adopted here included a detailed cochlear description (Verhulst et al., 2012; Altoè et al., 2014) and biophysical IHC-AN model (Altoè et al., 2018), but only included a phenomenological description of the CN and IC nuclei (Nelson and Carney, 2004). Even thought the same-frequency excitatory-inhibitory CN/IC model captures fundamental physiological observations (Nelson and Carney, 2004), a more detailed description for the brainstem/midbrain structures can be included in future modeling work. Another possible shortcoming of the model is seen when simulating absolute EFR_PT_ magnitudes. Even though there was a qualitative agreement between how SNHL affected the simulated and recorded responses, absolute EFR_PtN,PT_ magnitudes were smaller than simulated for both NH and HI subjects (Fig. 5a,d). This inconsistency likely stems from the extended contribution of off-CF cochlear regions which resulted in unrealistically high synchronized activity in the tails of the excitation patterns. Biological noise would have suppressed these off-CF contributions in the recordings, hence causing the mismatch in absolute magnitudes between recorded and simulated EFR_PT_s. The smaller experimental EFR_PtN,PT_ magnitudes also resulted in larger ABR_W-V_/EFR_P,PT_ ratios. These off-CF contributions did not affecting the simulated EFR_BB_s, which showed a good qualitative and quantitative match with the recordings.

Lastly, we constrained the simulations to two independent, but often coexisting, factors that can alter AEP characteristics: cochlear gain loss resulting from OHC damage, and synaptopathy reflecting IHC afferent synapse damage. There are other factors which may also alter the AEP (such as the medial olivocochlear reflex or middle-ear muscle reflex) and which were beyond the scope of this study. Understanding the influence of these other aspects on AEPs, can further help refine the sensitivity of proposed metrics.

### Near and supra-threshold cochlear amplification

Our study outcomes suggest the need to account for OHC integrity to make a precise synaptopathy quantification based on the supra-threshold EFR_PtN_ metric. However, cochlear amplification (and consequently its influence on supra-threshold AEPs) is lower for moderate-to-high sound levels than to low sound levels, as characterized by the BM input/output function at CF (Ren et al., 2011; Robles and Ruggero, 2001; Recio et al., 1998). Moreover, the relationship between near- and supra-threshold hearing impairment associated with OHC damage is not necessary straightforward due to individual variability of the stimulus-level dependent compressive growth of cochlear responses (Dorn et al., 2001; Neely et al., 2003; Schairer et al., 2006). Therefore, it is expected that, despite a significant relationship between the supra-threshold ABR_W-V_/EFR_P_ metric and audiometric thresholds, there is substantial individual variability between these two measures relationship (Fig. 8d,f). The observed variability of ABR_W-V_/EFR_P_ for a fixed audiometric threshold was greater than predicted by the model, because the model simulations did not account for individual differences in cochlear compression characteristics and applied a CF-independent OHC compression ratio which steepened as a function of cochlear gain loss (Verhulst et al., 2015, 2018). Furthermore, the supra-threshold ABR_W-V_/EFR_P_ metric has a broader frequency sensitivity (as compared to the adopted threshold metrics) and can hence be affected by the audiogram shape (Fig. 8a). The SNHL map concept is not bound by the supra-threshold ABR/EFR ratio and can also be constructed using the supra-threshold DPOAE magnitude, although DPOAEs should be recorded across the whole tonotopic range relevant to supra-threshold AEPs. It can be informative to quantify both the degree of near and supra-threshold cochlear gain loss (using the proposed ABR_W-V_/EFR_P,BW_ ratio) to determine the available dynamic range of hearing and to decide which hearing restoration strategies might be most successful in individual listeners.

### Translation of model insights and experimental evidence to clinical applications

We adopted an interdisciplinary approach to suggest the use of SNHL maps in the diagnosis of synaptopathy, which focused on stimuli which are commonly adopted in clinical practice and in animal studies of synaptopathy. In its present format, the proposed SNHL map provides a sensitive estimate of synaptopathy and supra-threshold OHC impairment with two stimulus conditions, yielding a 15-min recording. This offers a considerable benefit for its potential clinical uptake, especially given that EFRs and ABRs can already be recorded using clinical AEP devices. Another strong motivator of our approach is that other labs can map their previously collected ABR and EFR data onto a SNHL map to provide histological support and to obtain a more sensitive objective metric for synaptopathy which can be used to better understand individual differences in supra-threshold sound perception (e.g., Bharadwaj et al., 2015; Yeend et al., 2017; Prendergast et al., 2017a; Grose et al., 2017; Prendergast et al., 2017b; Guest et al., 2018; Valderrama et al., 2018).

However, it should be emphasized that this study is only a step towards a diagnostic paradigm for synaptopathy screening that can be used in a clinical context. Subsequent development to allow a translation into clinical practice requires rigorous testing with a large number of study participants with various degree of co-existing hearing pathologies to yield normative data and a precise continuum of the data distribution on the proposed SNHL maps. It may well be that preliminary stages of the clinical validation process will have an iterative character and will require further protocol adjustment or stimulus tuning to enhance the signal-to-noise ratio of the EFR metrics for measurement in clinical research.

## Data Availability

Data available from author upon reasonable request

## Acknowledgments

This work was supported by the German Research Foundation (DFG) Priority Program 1608 “Ultrafast and temporally precise information processing: Normal and dysfunctional hearing” VE924/1-1 (VV) and European Research Council (ERC) under the Horizon 2020 Research and Innovation Programme (grant agreement No 678120 RobSpear; SV).

## Author contributions

V.V. and S.V. designed research, V.V. and S.V. performed research, V.V. analyzed data; S.V and V.V. wrote the paper.

